# The Mediating Role of Structural Connectivity in Genetic Effects on Functional Brain Networks

**DOI:** 10.1101/2025.09.16.25335920

**Authors:** Simiao Gao, Shengxian Ding, Xinzhi Zhang, Zhiling Gu, Yize Zhao

## Abstract

Understanding the complex relationships among genetic variations, brain structural anatomy, and functional alterations is a fundamental yet challenging task in neuroimaging genetics. In this study, we employ a causal mediation framework under structural modeling to systematically investigate the mediating role of inter- and intra-network structural connectivity (SC) in linking whole-genome single nucleotide polymorphisms (SNPs) to brain functional connectivity (FC) across both resting-state and task-based conditions during neurodevelopment. Utilizing baseline and follow-up SC and FC network traits along with ∼500k SNPs along the genome from 11,666 unique subjects under the Adolescent Brain Cognitive Development (ABCD) study, we first conduct genome-wide association studies (GWAS) to identify candidate SNPs associated with structural and functional network traits. Subsequently, mediation analyses reveal key genetic exposures that directly influence brain functional networks and indirectly impact FC through SC network mediators. These results provide deeper insights into how genetic variations shape brain structural and functional network organizations, along with revealing the influence of brain anatomical topologies on functional fingerprints. This work enhances the understanding of the causal effect pathways among genetic factors and large-scale brain structural and functional networks, advancing our understanding of the genetic underpinnings of neurodevelopmental processes.

## 1 Introduction

Modern biomedical techniques have significantly advanced the study of human behavior. Advances in genotyping, in particular, have been playing a crucial role in uncovering the genetic underpinnings of behavioral traits including various complex psychiatric disorders (Bienvenu et al., 2011; Davies et al., 2011; Meyer-Lindenberg & Weinberger, 2006; L. Shen & Thompson, 2019). However, the mechanisms by which genetic variations shape these phenotypes remain largely unknown (Bi et al., 2017; T. Chen et al., 2022). This challenge is further compounded by the inherent complexity of cognitive and behavioral phenotypes, which are often self-reported and lack standardized definitions (Goldberg & Weinberger, 2004), making it more difficult to detect and interpret meaningful genetic associations.

Neuroimaging genetics has emerged as a prominent area of genetic research, aiming to bridge the gap between genetic variations and behavioral phenotypes (Bi et al., 2017; T. Chen et al., 2022; Jun et al., 2025), driven by the increasing availability of brain imaging, behavioral, and genetic data from large-scale studies. Specifically, neuroimaging traits serve as endophenotypes, offering a more direct representation of the biological mechanisms underlying psychiatric disorders and behavioral traits while maintaining a stable association with genetic variations. This facilitates more robust and interpretable insights into the genetic influences on cognitive abilities and psychiatric conditions, enhancing our understanding of their underlying etiology. Among various neuroimaging endophenotypes, functional connectivity (FC) and structural connectivity (SC) are of particular interest due to their characterization of the topological, network-level organization of brain structure and function (Friston, 1994; Park & Friston, 2013). FC measures the temporal correlations of neural activity across brain regions, typically using functional MRI (fMRI). It plays a crucial role in cognitive neuroscience by characterizing both intrinsic (resting-state) and task-based brain network dynamics. FC is often conceptualized in terms of functional networks or systems, where interconnected brain regions form subnetworks or modules to support cognitive, sensory, and motor functions (Yeo et al., 2011). In contrast, SC maps the anatomical connections between brain regions through white matter fiber tracks, primarily measured via diffusion imaging techniques. SC provides comprehensive anatomical underpinnings of white matter organization across parcellated brain regions, offering insights into how neural pathways facilitate communication. Critically, alterations of FC and SC networks have been linked to impairments in cognitive, sensory, and motor functions, as well as to various neurological and psychiatric disorders (Bullmore & Sporns, 2009; Stam, 2014). These findings suggest their essential roles in shaping neural function and behavior outcomes.

Previous studies have demonstrated significant associations between certain genetic variants and variations in cortical thickness and white matter microstructure (A. A. Joshi et al., 2011; Kochunov et al., 2011), highlighting the genetic underpinnings of brain structure. SC, which reflects the brain’s physical wiring, is shaped in part by genetic influences, as evidenced by heritability studies of white matter integrity and tract-based measures (B. Zhao et al., 2021). Similarly, FC is modulated by genetic factors, with genome-wide association studies (GWAS) and twin studies reporting moderate to high heritability of FC across large-scale brain networks (Yang et al., 2016; B. Zhao et al., 2022). These findings emphasize the importance of understanding how genetic variation contributes to both SC and FC, which together form the basis of cognitive and behavioral functions. Investigating their shared and distinct genetic architectures may provide insights into brain network organization and the genetic foundations of neurological and psychiatric conditions.

Research focusing on the relationship between SC and FC, commonly known as SC-FC coupling, has gained significant attention. This is largely because it offers valuable insights into how the brain’s structural framework shapes and underpins its functional dynamics. Zhang et al., 2011 laid the groundwork by demonstrating the first evidence of significant correlations between SC and FC, suggesting that structural pathways are key determinants of functional organizations. Sun et al., 2017 extended this work by investigating the variability of SC-FC coupling across individuals, highlighting how individual differences in white matter integrity influence functional network topology. Cao et al., 2020 explored how SC-FC coupling changes across different cognitive states, emphasizing the dynamic nature of brain networks and their reliance on underlying structural substrates. Meanwhile, by measuring SC-FC coupling as a unified phenotype, heritability and genetic association studies (Dai et al., 2024; Gu et al., 2021) have begun to be conducted in the young adult population to characterize the genetic contributions on the shared interplay of SC and FC. These findings suggest the complex interplay between genetics and brain connectivity, as well as the intricate relationship between structural and functional connectivity. They also highlight the need for a deeper investigation into the causal mechanisms linking genetic factors, SC variations, and FC alterations.

Mediation analysis is a statistical framework to reveal casual effect pathways between an exposure and an outcome potentially mediated by a mediator. In the literature of neuroimaging genetic studies using mediation frameworks, researchers have focused primarily on elucidating how genetic effects on cognitive phenotypes are mediated by specific brain functions. For example, Z. Chen et al., 2024 explored the role of brain imaging phenotypes in mediating the relationship between genetic variations and cognitive outcomes, with a particular focus on aging and drug targets. Additionally, Mu et al., 2024 utilized genome-wide mediation analysis to investigate how genetic influences on amyloid and tau pathology contribute to cognitive decline through specific brain imaging phenotypes in Alzheimer’s disease. Despite these advances, a critical gap remains in understanding how genetic effects on functional networks are mediated by structural anatomical regions, highlighting the need for a more integrated approach to studying brain connectivity and its interconnections. In addition, while previous studies have provided valuable insights into genetic influences on connectivity within individual resting-state networks, such as the default mode network or the salience network (Tissink et al., 2023), the genetic basis of connectivity between different functional networks remains largely unexplored. Moreover, most existing research has focused on resting-state FC, while relatively little is known about how genetic variants influence connectivity patterns during task-related activity. A deeper understanding of these processes requires investigating how the brain’s anatomical structure shapes FC across both resting-state and task-based conditions.

To fill this gap, we adopted an analytical framework to elucidate the pathways through which genetic variants influence functional network-based connectivity in both resting-state and task-related activity, mediated by structural intra- and inter-regional connectivity. In this framework, we explicitly considered *brain structural networks as mediators and brain functional networks as outcomes*, providing a comprehensive view of the interplay between genetic variants, structural connectivity, and functional network organization. In particular, we first conducted GWAS to identify genetic variants associated with various neuroimaging measures, encompassing both structural and functional network traits. Then, we performed mediation analysis on each significant single nucleotide polymorphism (SNP) identified in the GWAS, to investigate how SC - both within and between brain regions - mediates the effects of genetic variants on FC. Specifically, we examined how structural intra- and inter-regional connectivity shapes functional intra- and inter-network connectivity, thereby elucidating the pathways through which genetic factors influence brain function. By integrating structural and functional pathways within a unified mediation framework, we gained deeper insights into the genetic mechanisms underlying brain network organization.

For this study, we investigated the genetic architecture underlying SC and FC networks in preadolescents from the Adolescent Brain Cognitive Development (ABCD) study (11,666 participants). It provides multi-modal neuroimaging, genetics, cognitive, and behavioral measures at baseline and two-year follow-up, which enables in-depth investigations into the genetic basis of neurodevelopmental processes. The availability of two time points enables us to evaluate the static genetic influences across time on brain network organization. We performed genetic association analysis between whole genome SNPs and SC and FC network traits at both time points, and assessed the replication of genetic-trait associations by examining overlapping genomic regions mapped to cytogenetic bands. Through mediation analysis with leading SNP exposures, we quantified the indirect effect of SC in mediating genetic influences on FC at both time points and assessed the consistency of the implicated genomic regions, SC traits, and FC network alterations. Our findings revealed multiple mediation pathways through SC network traits, linking causal genetic signals to FC networks. Notably, some of these pathways involved SC traits spatially distant from the corresponding FC traits, suggesting that genetic influences on brain connectivity can extend beyond anatomically proximal regions. Furthermore, the moderate to high replication rates of SC-mediated pathways connecting SNPs to FC traits across both time points indicate the stability of certain genetic effects on brain connectivity throughout adolescent development.

The rest of this article is organized as follows. Section 2 details the dataset, neuroimaging data preprocessing, derivation of functional and structural network traits, GWAS procedures, mediation analyses, and effect pathway quantification. This section also introduces consistency evaluations, including functional annotation, identification of shared loci for causal SNPs, and replication of mediation pathways using the two-year follow-up data. Section 3 presents our primary baseline findings along with the consistency evaluations on the two-year follow-up data, followed by a discussion of potential directions for future research in Section 4.

## 2 Methods and Materials

### 2.1 The ABCD study

Our study samples come from this landmark ongoing children’s study, the largest prospective research project that collects a wealth of measures including multi-modal neuroimaging, genetics, cognitive, behavioral, and youth and parent self-report metrics among preadolescents, with the general goal to understand how biological and environmental factors influence brain development from childhood to young adulthood. In particular, we utilized the ABCD Data Release 5.1, which involves 12,000 early elementary school children recruited from 21 sites across the United States (Garavan et al., 2018), providing minimally processed neuroimaging data that contains initial data (baseline) on the full participant cohort, and a second imaging timepoint after two years (two-year follow-up). The data are stored in the NIMH Data Archive (NDA) (https://nda.nih.gov/abcd).

#### 2.1.1 Multi-modal Imaging

We took advantage of the multi-modality data within the ABCD study, which provides high-resolution structural MRI (sMRI), advanced diffusion MRI (dMRI), resting-state fMRI, and task-based fMRI. The dMRI and sMRI were used to recover SC, and the resting-state fMRI, and task-based fMRI were used to derive FC.

The ABCD neuroimaging protocol begins with a localizer scan to position the participant’s head, followed by a high-resolution 3D T1-weighted sMRI to capture detailed brain anatomy. Next, resting-state fMRI is acquired during two periods of quiet rest (eyes open, fixating on a crosshair). Subsequently, dMRI is collected to map white matter pathways by tracking water diffusion patterns. A T2-weighted scan is then acquired for complementary anatomical contrast, and brief field maps are obtained to correct geometric distortions in functional and diffusion data. The session concluded with one to two additional resting-state fMRI runs and task-based fMRI scans. Specifically, the resting-state fMRI are recorded when participants remain still to assess intrinsic brain connectivity. The task-based fMRI involves cognitive and emotional tasks such as the Monetary Incentive Delay (MID) task for reward processing (Knutson et al., 2000), the N-Back (nBack) task for working memory (Cohen et al., 2016), and the Stop Signal Task (SST) for response inhibition and cognitive control (Logan, 1994). Standardized across all sites, the protocol prioritizes motion mitigation (e.g., head stabilization, mock scanner training) and harmonized 3T scanner settings to ensure consistency in tracking longitudinal brain development in children and adolescents (Casey et al., 2018). Detailed information on the multi-modal imaging data can be accessed at https://wiki.abcdstudy.org/release-notes/imaging/overview.html.

Moreover, we conducted Quality Control (QC) as follows: For task-based fMRI, only participants with a mean framewise displacement (FD) below 0.15 mm were included. For resting-state fMRI, we analyzed one scan per participant, selecting the run with the lowest FD when multiple scans met the threshold. Additionally, participants with low-quality anatomical images, as flagged by FreeSurfer (ABCD NDA name: fsqc_qc), were excluded to ensure data quality.

#### 2.1.2 Genotype data

The recent ABCD Release 5.1 includes both pre- and post-imputed genotype data for 11,666 unique participants. The imputed genotype data, derived from the TOPMed reference panel, encompassed approximately 300 million variants. To account for population stratification, we conducted QC on the pre-imputed genotype data (∼ 500,000 SNPs) using the RICOPILI pipeline (Lam et al., 2020) with stringent filtering criteria. SNPs were excluded if they had a minor allele frequency (MAF) below 0.01, failed the Hardy-Weinberg equilibrium (HWE) test (*p <* 1 × 10^−6^), or did not meet the linkage disequilibrium (LD) pruning threshold (200/50/0.25). Genetic relatedness was assessed using kinship coefficients, and only unrelated individuals (kinship *<* 0.125) were retained. Principal component analysis (PCA) was performed on the QC-filtered genotype data to estimate population structure and infer European genetic ancestry among participants (see Figure 1). To ensure a genetically homogeneous population group, only individuals who self-identified as “white” were included in subsequent analyses. Population outliers were removed by retaining individuals whose scores on the first two principal components fell within ±3 standard deviations (SD) from the group mean (Teeuw et al., 2023). Participants were included if there was no evidence of sex mismatch between phenotypic and genetically determined sex, genotype missingness was below 1%, and autosomal heterozygosity deviation fell within ±3 SD. After this procedure, 4,657 participants were retained.

**Figure 1:**
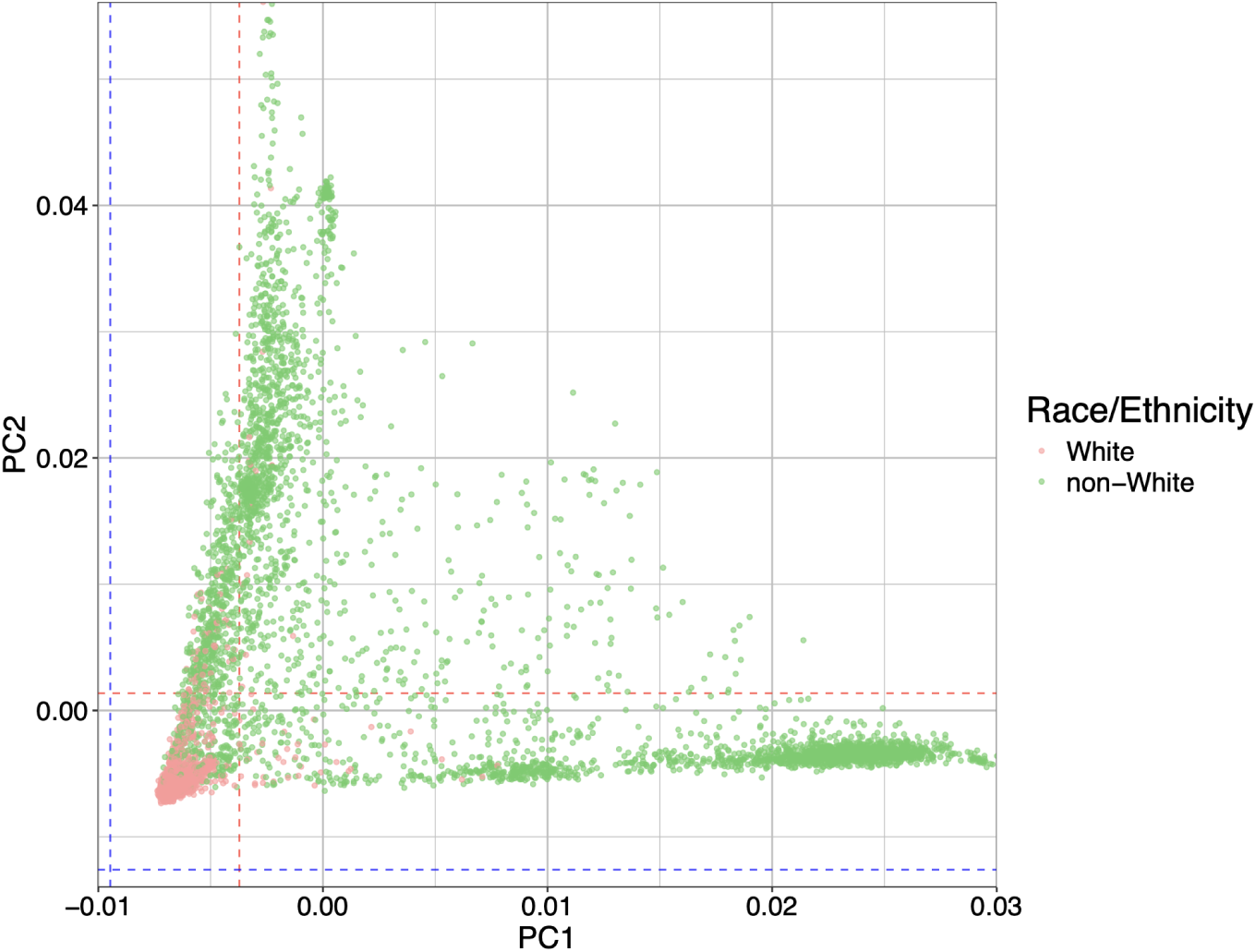
First two principal components (PC1 and PC2) for European genetic ancestry.

Subsequently, we performed post-imputation QC to exclude SNPs with low MAF (< 0.01), poor imputation quality (INFO < 0.8), or failure to meet the HWE threshold (*p <* 1 × 10^−6^). After these filtering steps, approximately 650,000 autosomal SNPs were retained for the subsequent GWAS analysis.

#### 2.1.3 Covariates and confounders

To ensure robust and reliable results, we incorporated a comprehensive set of covariates to account for potential confounding factors that could influence genetic associations. These covariates encompassed age, sex, and the top ten principal components (PCs) derived from SNP data to adjust for population stratification. Additionally, we controlled for socioeconomic and environmental influences by including combined family income and parental education. Puberty status was considered to account for developmental differences, while study site was included to control for site-specific variability. We also adjusted for handedness, given its known association with brain function, and intracranial volume to account for individual differences in brain size.

We applied the same covariate adjustments in both GWAS and mediation analyses. After aligning subjects with European genetic ancestry, complete confounder information, and all MRI modalities (dMRI, sMRI, and fMRI), we obtained different sample sizes for each fMRI state. At baseline, the final set included 2,447 subjects for resting-state fMRI, 1,804 for the MID task, 1,464 for the nBack task, and 1,768 for the SST. The same processing pipeline was applied to the two-year follow-up data, resulting in 966 subjects for resting-state fMRI, 581 for the MID task, 488 for the nBack task, and 573 for the SST.

### 2.2 Construction of FC network traits

Following the procedure outlined in Wang et al., 2024, we then constructed FC matrices for each participant using raw DICOM image data from resting-state and three cognitive task fMRI sessions. Pre-processing was conducted with BioImage Suite (A. Joshi et al., 2011) following standard protocols, including motion correction, registration to MNI space, and anatomical parcellation using a 268-node whole-brain atlas (X. Shen et al., 2013). To reduce noise, covariates such as linear, quadratic, and cubic drifts, 24-motion parameters (Satterthwaite et al., 2013), cerebrospinal fluid signal, white matter signal, and global signal were regressed out; after that, the data was temporally smoothed using a Gaussian filter (*σ*=1.95). FC matrices were then generated for each state and participant by computing Pearson correlation coefficients between the time courses of all node pairs, followed by Fisher z-transformation.

To explore the relationship between SNPs and functional networks, and to enhance result interpretation, the 268 × 268 FC matrices were further organized into network-based parcellations. Specifically, the Shen 268-node atlas (X. Shen et al., 2013) was used to divide the whole brain into 268 distinct regions of interest. These regions were further grouped into eight large-scale functional networks (Medial frontal, Frontoparietal, Default mode, Subcortical – cerebellum, Motor, Visual I, Visual II, and Visual association), reflecting different functional systems that capture the complexity of neural interactions. This network-based parcellation provided a structured framework for examining both intra-network and inter-network FC dynamics across the brain. This process is illustrated in Figure 2, yielding 36 functional network traits. A full list of functional network traits utilized in the analyses is provided in Supplemental Table S1.

**Figure 2:**
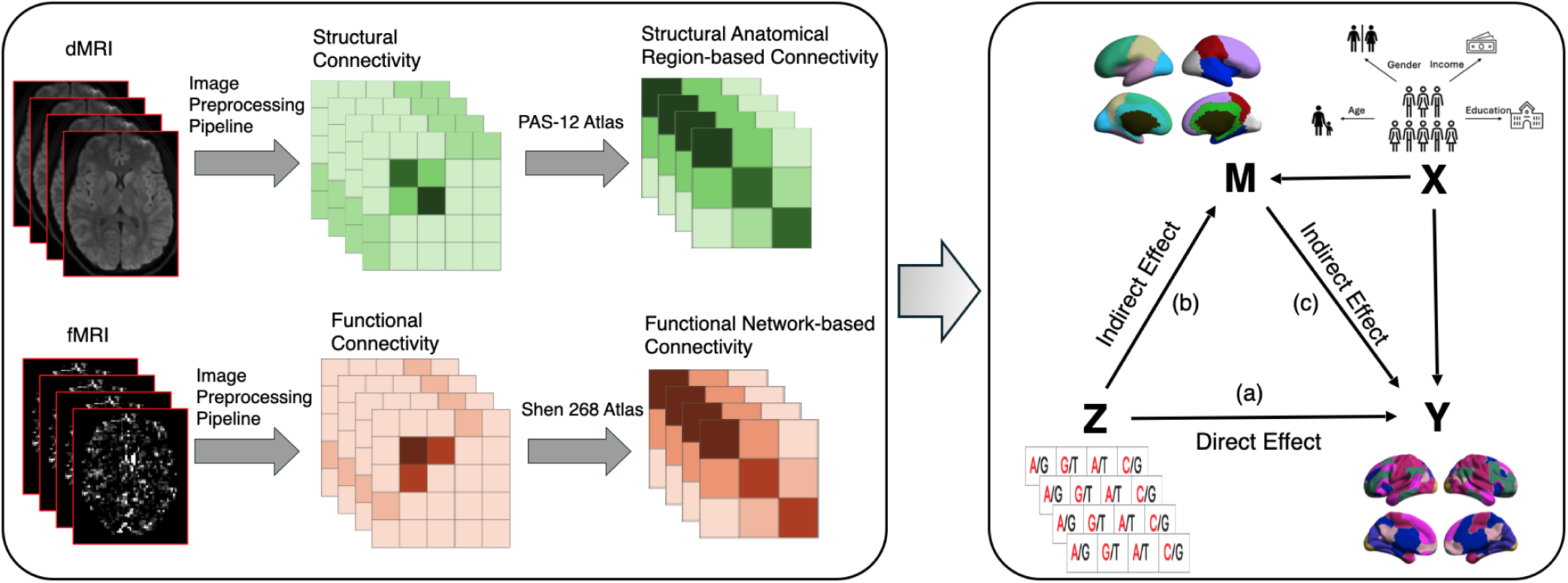
Overview of the FC and SC network traits generation process and the downstream mediation framework. (1) Resting-state and task-based fMRI scans, along with dMRI scans, undergo image preprocessing as described in Section 2.2. (2) High-dimensional functional and structural connectivity matrices are constructed from the preprocessed MRI data for each participant. (3) These matrices are then organized into network-based parcellations for FC and anatomically meaningful parcellations for SC, from which the intra- and inter-network SC and FC traits are extracted. (4) GWAS analysis is performed in (a) and (b) to identify SNPs that are significantly associated with the FC and SC traits. (5) Using GWAS-identified SNPs as exposure (*Z*), mediation analysis is conducted with SC and FC traits, which serve as outcomes (*Y*) and mediators (*M*) adjusting confounders (*X*). (6) Significant indirect effects, as indicated in (b) and (c), are examined to uncover the intermediate genetic influences on functional subnetworks.

### 2.3 Construction of SC network traits

To avoid arbitrarily choosing atlases, SC matrices were generated using the Surface-Based Connectivity Integration (SBCI) pipeline (Cole et al., 2021). This approach uses the white surface (the interface between white and gray matter) to construct SC without being constrained by predefined brain parcellations (Dai et al., 2024). We reconstructed SC using dMRI post-eddy-current correction and T1-weighted (T1w) images after gradient correction. Specifically, minimally preprocessed dMRI data were obtained from data repositories, where preprocessing included *b*_0_ intensity normalization, susceptibility-induced distortion correction, motion correction, and eddy current correction, as detailed in Glasser et al., 2013. T1w anatomical images were registered using ANTs, followed by surface reconstruction using the recon_all tool from FreeSurfer (http://freesurfer.net/). Furthermore, the tractography algorithm SET (St-Onge et al., 2018) was employed to extend connectivity data to a 32k white surface, ensuring high-quality SC reconstruction for downstream analyses. The final output consisted of the 5124 × 5124 SC matrices, which were used for downstream analyses.

To enhance the interpretability of results, the high-dimensional SC matrices were structured into anatomically meaningful subnetworks using the PALS-B12 atlas (Van Essen, 2005). This atlas defines ten distinct cortical regions, including the frontal, parietal, limbic, temporal, and occipital lobes, each mapped separately for the left and right hemispheres, while excluding the wall region. The process of constructing the SC network traits is illustrated in Figure 2. A full list of the resulting structural network traits is provided in Supplemental Table S2.

### 2.4 Genome-Wide Association Studies (GWAS)

In mediation analysis, identifying a significant causal mediation pathway requires significant associations between the genetic variants (exposures) and both the mediators (SC network traits) and outcomes (FC network traits). Therefore, GWAS serves as a critical initial screening step to identify potential genetic exposures by detecting SNPs significantly associated with the derived functional and structural network traits for each brain state.

GWAS was performed via PLINK (Purcell et al., 2007) with all the relatives removed, where age, sex, the top 10 genetic PCs, family income, parental education, puberty status, study site, handedness, and intracranial volume were adjusted for. The phenotypes analyzed included the constructed FC network traits and SC network traits. Following quality control, approximately 650,000 SNPs remained in the genotype data. To refine the candidate set of causal SNPs with potential mediation effects through the SC mediators on the FC network traits, SNPs with high p-values were filtered out after applying a False discovery rate (FDR) correction to control for multiple testing. The remaining candidate SNPs that passed the FDR-adjusted significance threshold (*p <* 0.05) were subsequently examined in mediation analysis to evaluate their potential indirect effects on the FC network traits through SC mediators.

### 2.5 Mediation analyses for brain network genetics

Mediation analysis is a widely used approach to estimate causal relationships, providing insight into how a third variable, referred to as mediators or mediating variables, influences the effect of an independent variable on a response variable. In this work, we hypothesize that genetic variants impact FC network alterations both directly and indirectly through their effects on SC network architectures. Thus we employ a mediation framework to investigate the causal effect pathways among genetic factors, intra-and inter-network SC and FC traits. In this framework, SC network traits (*m*) act as potential mediators between genetic variants (*z*) and FC network traits (*y*) with the influence of SC on FC networks being partially shaped by genetic contributions. By further incorporating covariate vectors ***x***, including an intercept term to account for confounders, we propose the following mediation framework:

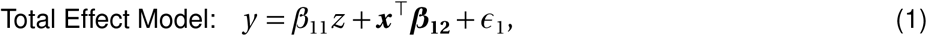

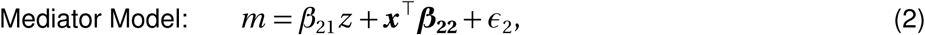

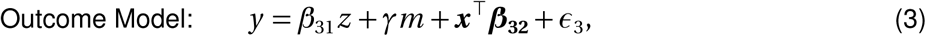

where the random errors *ε*_1_, *ε*_2_ and *ε*_3_ are assumed to be independent and follow normal distributions. Here, *β*_11_ relates genetic variate (exposure) to the FC network trait (outcome), *β*_22_ captures the correspondence between genetic variate (exposure) with the SC network trait (mediator), and *β*_31_ and *γ* characterizes the effect of the exposure on the outcome and the effect of the mediator on the outcome, respectively. Following the counterfactual framework concerning mediation in the causal inference literature, we denote *M_i_* (*z*) and *Y_i_* (*z*, *M_i_* (*z*)) as the potential mediator and outcome under the exposure *Z_i_* = *z*, and *Y_i_* (*z*, *m*) as the potential outcome under *Z_i_* = *z* and *M_i_* = *m* . Additionally, the following assumptions known as the sequential ignorability assumptions (VanderWeele & Vansteelandt, 2009) are also assumed to provide identification for the casual effects, including 1) *Y_i_* (*z*, *m*) ⊥ *Z_i_* | *X_i_*, i.e., no unmeasured confounders for the SNP-functional trait relationship; 2) *Y_i_* (*z*, *m*) ⊥ *M_i_* | *Z_i_*, *X_i_*, i.e., no unmeasured confounders for the structural trait-functional trait relationship; 3) *M_i_* (*z*) ⊥ *Z_i_* | *X_i_*, i.e., no unmeasured confounders for the SNP-structural trait relationship; 4) *Y_i_* (*z*, *m*) ⊥ *M_i_* (*z* ^∗^) | *X_i_* for all levels of *z*, *z* ^∗^ and *m*, i.e., no SNP-induced confounding for the structural trait-functional trait relationship. These assumptions serve as the standard requirements for a causal mediation paradigm and have been extensively discussed and adopted in the latest neuroimaging mediation studies (Lindquist, 2012; Tian et al., 2024; Y. Zhao et al., 2022).

### 2.6 Effect pathways and mediating role of SC on FC

Based on the above structural modeling, we can quantify the effect pathways among each genotype, SC network trait and FC network trait through the natural direct effect (NDE), the natural indirect effect (NIE) and the total effect (TE) given two arbitrary levels of the exposure *Z_i_* = *z* and *z* ^∗^ as follows:

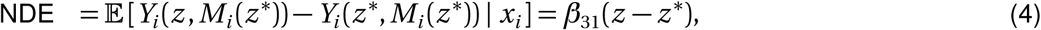

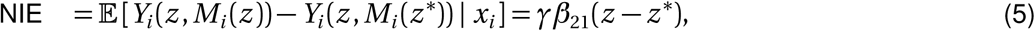

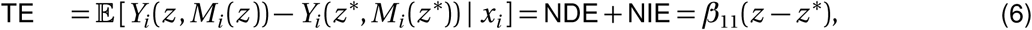

where Equation (6) demonstrates that the TE of the genetic variant *z* on the FC network trait *y* can be decomposed into the summation of NDE in (4) and NIE in (5). To determine whether a genetic variant influences FC indirectly through SC mediator, we focused on identifying significant mediation pathways, i.e., *z* → *m* → *y*, when NDE was statistically significant. We estimated NDE and NIE for each potential mediation pathway, along with their uncertainty quantification. To characterize the mediation effect size, we computed the mediation proportion (MP) (Lee et al., 2024) as

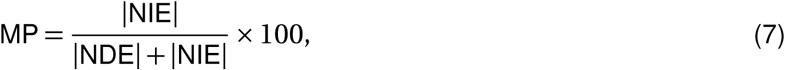

which is the proportion of the TE mediated through the SC mediator. Absolute values were used in this calculation to account for situations where the direct and indirect effects had opposite directions, a scenario known as inconsistent mediation(Coutts & Hayes, 2023). In such cases, the effects could otherwise offset each other, leading to an underestimation of the mediated effect (Alwin & Hauser, 1975). To adjust for multiple hypothesis testing in mediation analysis, FDR correction was applied to account for the number of mediation tests conducted, and a significance threshold of 0.05 was adopted. Significant indirect effects were thus identified in the mediation analysis, ensuring that only statistically robust pathways were retained.

Furthermore, we explored the mediating role of structural networks in shaping both intra-network and internetwork FC. In neurological disorders, SC and FC dynamics often involve cross-network interactions rather than being restricted to co-localized networks. Structural disruptions, such as focal lesions, can trigger functional alterations that propagate through polysynaptic pathways, resulting in functional connectivity changes in distal brain regions that lack direct structural connections. Therefore, the coupling between structural and functional networks of interest was examined to better understand this relationship. By investigating how structural pathways transmit genetic effects on functional brain organization, we aimed to shed light on the mechanisms through which genetic variation influences large-scale brain network dynamics and contributes to cognitive functions, neurodevelopment, and susceptibility to neurological disorders.

### 2.7 Functional annotation and shared loci for causal genetic factors

To better understand the biological interpretation of the identified causal SNPs, we employed Functional Mapping and Annotation (FUMA) for functional annotation. FUMA is an online platform designed to functionally map and annotate SNPs identified as significant in GWAS (Watanabe et al., 2017). GWAS summary statistics from Equations (1) and (2) were uploaded to FUMA for analysis. Annotation was performed using ANNOVAR, and genomic loci were defined by selecting all independent significant SNPs with *r* ^2^ *<* 0.1 and LD blocks within 250 kb, using the 1000G Phase3 EUR reference panel. Within each locus, SNPs were assigned to genes using both positional mapping and expression quantitative trait loci (eQTL) mapping. Positional mapping was performed by linking SNPs to nearby genes within a 1-Mb window based on known genomic associations. eQTL mapping leveraged brain tissue expression data from well-established repositories, including GTEx (Consortium et al., 2015) and BRAINEAC (Ramasamy et al., 2014), to identify regulatory relationships between genetic variants and gene expression. The mapped genes were analyzed to uncover potential links to brain development or the dysfunction of functional systems. Additionally, we examined eQTLs, which reflect the molecular regulation of gene expression across different brain regions, providing further insights into the biological mechanisms underlying genetic influences on neuroimaging traits.

In addition to utilizing the FUMA platform, we mapped the loci of GWAS-identified SNPs to their corresponding cytogenetic bands to better understand their genomic distribution. Cytogenetic bands, defined by chromosomal staining patterns, offer an additional layer of genomic annotation that complements sequence-based mapping (Schneider et al., 2017). Cytogenetic mapping provides a structural context for SNPs, allowing us to examine their localization within chromosomal regions characterized by structural variations, gene densities, and known disease-associated loci (Hurles et al., 2008). This approach facilitates the identification of potential functional hotspots and the prioritization of SNPs for further validation and biological interpretation. Hence, the results were presented at the cytoband level to enhance the resolution of GWAS findings and uncover mechanistic insights into observed genetic associations. Moreover, in the mediation analysis, this refinement highlighted genomic regions associated with mediated effects on brain connectivity.

### 2.8 Consistency evaluation on two-year follow-up data

Previous research has shown that certain genetic influences remain relatively static across time while others may specifically impact brain development (Brouwer et al., 2022). These stable influences form the foundation for consistent network architecture. By prioritizing stable signals, our analysis aimed to highlight genetic variants with enduring effects on SC and FC network traits. Identifying such stable genetic signals is crucial, as they may reflect fundamental biological mechanisms underpinning brain connectivity that persist despite ongoing neurodevelopmental changes. To assess the robustness and reproducibility of the identified genetic associations and mediation pathways, we leveraged the two-year follow-up data, which provided an opportunity to assess temporal stability and the potential influence of neurodevelopmental changes on SC and FC. Of the participants included in the baseline dataset, 966 shared subjects were available for follow-up. While SC and FC may evolve during adolescence, observing consistent genetic signals across time points strengthens the evidence for stable genetic influences on brain network organization.

The replication analyses consisted of two main steps:

**(1) Replication of GWAS findings:** We repeated the GWAS in the two-year follow-up data using the same statistical models and covariate adjustments described in Section 2.4. We conducted functional annotation on significant SNPs after FDR correction and compared the results to the baseline data. Reproducibility was assessed by examining overlapping genomic regions mapped to cytogenetic bands. Despite potential temporal changes in SC and FC during this neurodevelopmental stage, observing concordant genomic regions provides evidence for stable, trait-relevant genetic influences.
**(2) Evaluation of network-level mediation consistency:** For SNPs significantly associated with SC and FC network traits, we evaluated their roles in the brain network genetics mediation analysis (Section 2.5). Specifically, we examined mediation pathways with statistically significant NIE, assessing consistency of the genomic regions and the implicated SC and FC network traits across datasets. Consistent network-level mediation findings further support the robustness of the identified genetic influences and their potential biological relevance.

Together, these replication analyses underscored the reliability of our results and highlighted genetic variants that exerted stable influences on SC and FC network traits, despite ongoing neurodevelopmental changes.

## 3 Results

### 3.1 Genetic associations for SC and FC network traits

As described in Section 2.4, GWAS were performed to investigate the associations between individual SNPs and each SC and FC network trait. For the baseline ABCD data, after controlling for an expected FDR of 0.05 for multiple testing, we identified 3,255 significant associations between SNPs and the derived SC and FC network traits in the resting-state condition, which reflects intrinsic neural activity. These associations involved 1,680 SNPs distributed across 203 genomic regions (cytogenetic bands) and were linked to 10 SC and 19 FC network traits. For the MID task, which assesses reward processing, 207 significant associations were identified, involving 199 SNPs spanning 71 genomic regions and associated with 16 SC and 6 FC network traits. For the SST, which evaluates response inhibition, we observed 175 significant associations, with 167 SNPs mapped to 72 genomic regions and linked to 9 SC and 13 FC network traits. Finally, for the nBack task, which examines working memory, 269 significant associations were detected, comprising 231 SNPs distributed across 97 genomic regions and associated with 17 SC and 7 FC network traits. These results highlighted distinct genetic influences on brain anatomical pathways as well as interconnected neural activities during the intrinsic resting-state and task-related cognitive processing.

We then analyzed the GWAS results using FUMA to further identify and understand the biological bases for the lead SNPs, following the settings outlined in Section 2.7. Specifically, we identified 220 independent lead SNPs for resting-state FC network traits, 12 for the MID task, 25 for the SST, and 46 for the nBack task. Functional and structural networks significantly associated with SNPs identified in the GWAS analysis are presented in Table 1, where the color gradient represents the number of SNPs linked to each network. In terms of FC, the Medial frontal functional network emerged as a key region linked to SNPs with the smallest GWAS p-values in the resting-state condition. Additionally, significant SNP associations were identified for task-based FC, including the SST and nBack tasks. These associations were most prominent in FC between the Medial frontal and Visual I networks, suggesting genetic influences on the integration of high-order executive and visual processing regions. Similarly, significant SNPs were linked to connectivity between the Frontoparietal and Motor networks, highlighting potential genetic modulation of cognitive-motor interactions during task performance. For SC, genetic associations were consistently observed in the frontal and limbic lobes of the right hemisphere, suggesting a strong genetic contribution to the anatomical connectivity of these regions, which are crucial for executive function, emotion regulation, and memory processing. Subsequently, these significant SNPs served as candidate SNPs in mediation analysis for assessing the potential mediation effect on the FC network traits through SC mediators.

**Table 1:**
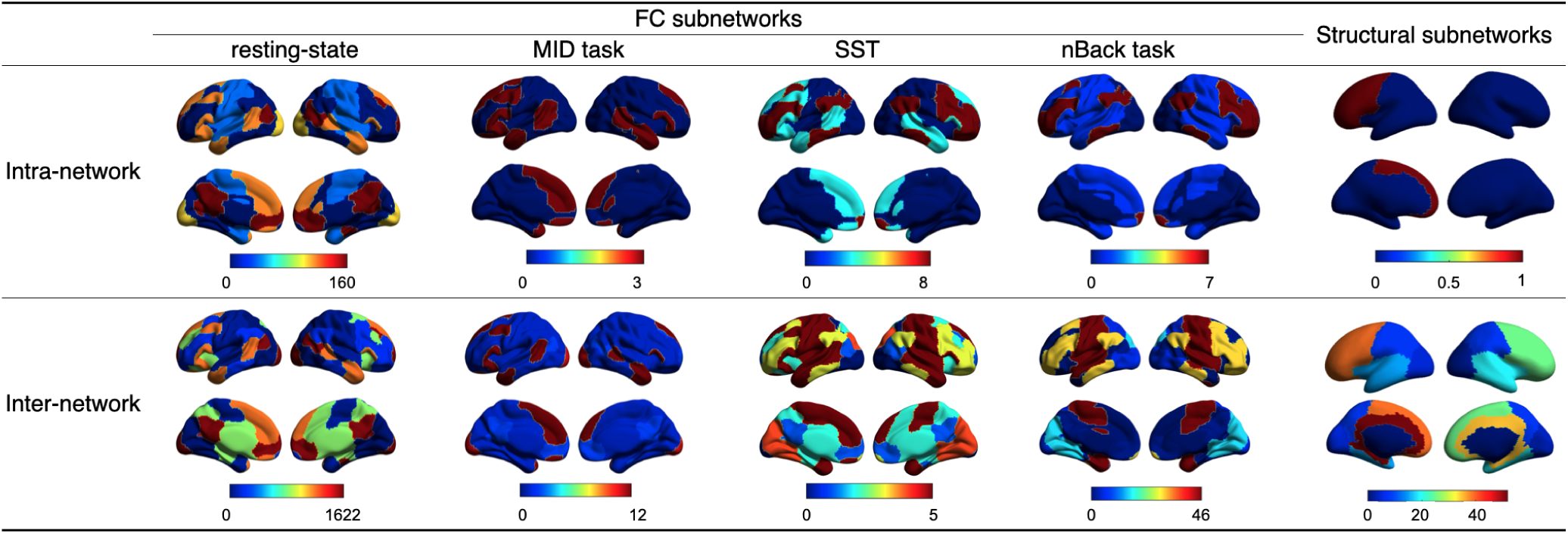
Functional and structural networks significantly associated with SNPs identified in the GWAS analysis, with the color gradient representing the number of SNPs linked to intra- and inter-network traits.

#### 3.1.1 Replication of genetic associations on two-year follow-up data

To assess the robustness of the genetic associations between SNPs and neuroimaging traits, we conducted replication analyses using the baseline and the two-year follow-up data. In particular, we examined the overlapped genomic regions identified in both datasets.

At baseline, for resting-state, we identified 284 associations involving 282 SNPs mapped to 91 genomic regions and linked to 18 network traits (13 SC and 5 FC). For the MID task, 403 associations were observed, with 341 SNPs spanning 118 genomic regions and associated with 36 network traits (23 SC and 13 FC). For the SST, 374 associations were identified, involving 345 SNPs distributed across 126 genomic regions and linked to 16 SC and 10 FC network traits. In the nBack task, 162 associations were detected, comprising 161 SNPs across 72 genomic regions and associated with 11 SC and 1 FC network traits.

The two-year follow-up data produced 81 genomic regions that overlapped with the baseline for the restingstate condition, 49 for the MID task, 56 for the SST, and 43 for the nBack task. Figure 3 presents the genomic regions associated with GWAS-identified SNPs that passed the initial screening and were potentially linked to network traits for the MID task. The ideograms corresponding to SST and nBack tasks are shown in Supplemental Figure. S1 and Supplemental Figure. S2. The resting-state condition is not shown due to the large number of associations identified. Specifically, Figure 3 conveys a wealth of information that across different functional states, several network traits were identified at specific cytobands, with varying degrees of replication in the two-year follow-up dataset. For the resting-state, we detected 13 network traits at q21.1 in the baseline dataset, while the two-year follow-up dataset revealed 3 network traits at the same cytoband, with one overlapping with the baseline set. In the MID task, two SC network traits were associated with q32.1, with one replicated in the two-year follow-up set. Similarly, for the SST, four network traits were linked to q34.1, of which one was replicated in the two-year follow-up analysis. Lastly, for the nBack task, two network traits were identified at baseline, and both were consistently observed in the two-year follow-up dataset, indicating stronger reproducibility in this case.

**Figure 3:**
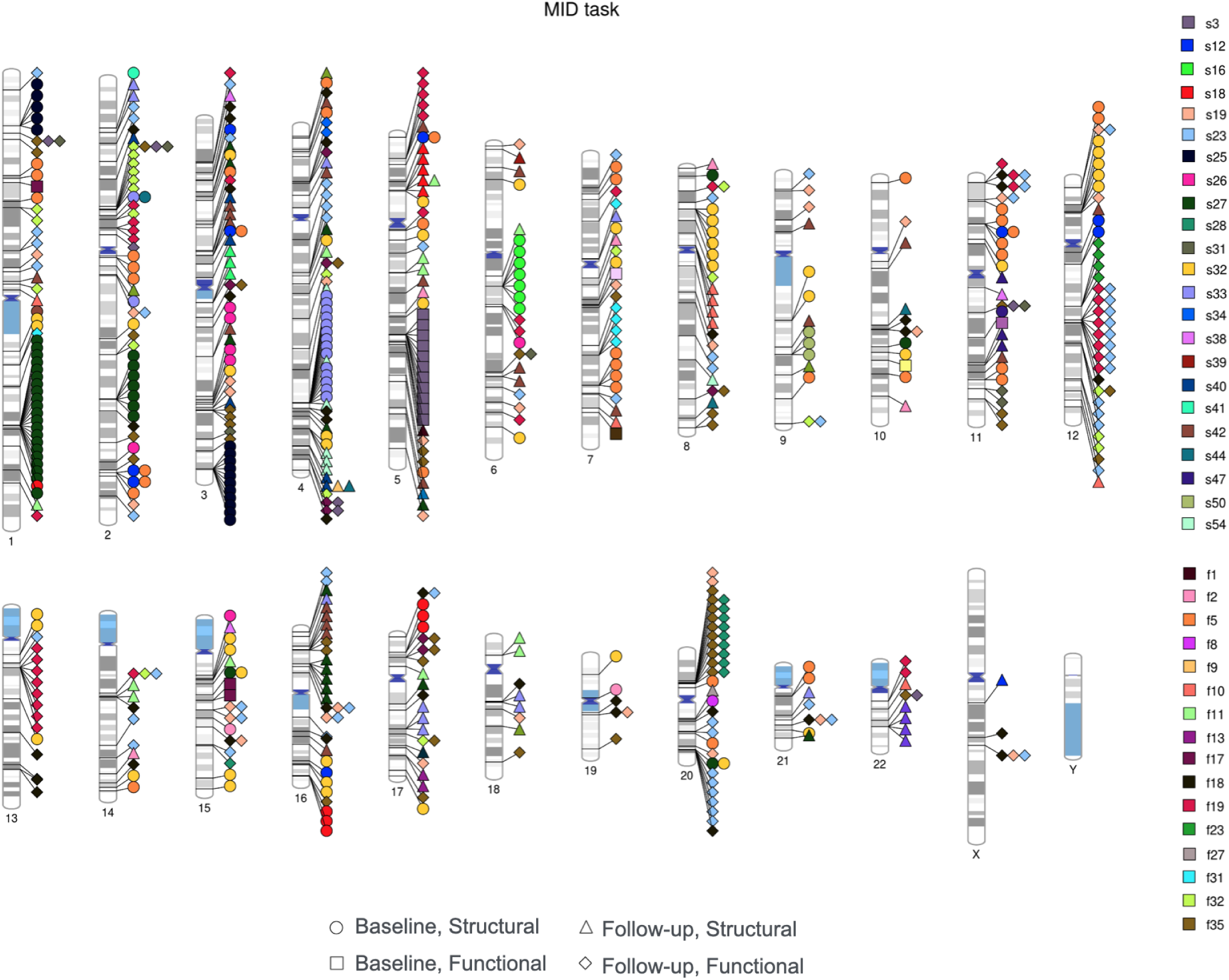
Ideogram of genomic regions influencing FC and SC network traits. Colors distinguish between intra- and inter-network connectivity for both functional and structural modalities, while the shape of the points represents the dataset to which they belong. Each individual point signifies that one network trait is associated with an SNP located within the corresponding genomic region. pathway from genetics to FC has undergone a transition from initial observation to the two-year followup. It is important to note that MP was calculated based on the absolute value of effects without accounting for effect directionality; see Equation (7).

Overall, the results indicated a moderate to high degree of reproducibility across datasets spanning two years. The identified overlapping genomic regions offer valuable insights into the static associations between genetics and both structural and functional brain connectivity, and potentially common genetic mechanisms. The number of overlapping genomic regions varied across different functional states, suggesting potential state-specific genetic influences on network traits. And the overall difference between baseline and follow-up may reflect the refinement of the brain network over time.

### 3.2 SC as mediators of genetic influences on FC

We further explored the role of SC network traits in mediating the impact of causal SNPs on FC network traits (significant indirect effect pathways are provided in the Supplementary Materials). This could comprehensively uncover all the potential effect pathways across key genetic factors, inter- and intra-network traits generated from whole brain SC and FC.

#### 3.2.1 Mediation pathways via SC network traits

For the resting-state condition, a total of 4,556 mediation pathways were identified, involving 658 SNPs mapped to 145 cytobands, with 51 SC mediators and 35 FC network traits. In the MID task, 465 pathways were detected, comprising 76 SNPs located on 32 cytobands, with 34 SC mediators and 32 FC network traits. For the SST, 677 mediation pathways were identified, including 97 SNPs across 45 cytobands, with 39 SC mediators and 33 FC network traits. Lastly, in the nBack task, 918 pathways were observed, involving 91 SNPs distributed across 51 cytobands, with 35 SC mediators and 28 FC network traits. Figure 4 illustrates the number of cytoband-level mediation pathways associated with each FC network trait, with SC network traits serving as mediators. Due to the large number of mediation pathways identified in the resting-state condition within the baseline set, pathway counts for each combination of FC and SC network traits exceed those of the three task-based states. In contrast, the number of pathways showed minimal variation among the MID task, SST, and nBack task conditions. Figure 5 displays boxplots of MP - the proportion of the SNP effect on FC network traits mediated by SC network traits - across the four cognitive states and two time points, with MP ranging from approximately 22% to 40%. Notably, the Mann-Whitney U tests revealed significant differences between the baseline and two-year follow-up sets for the resting-state and MID task conditions, indicating the

**Figure 4:**
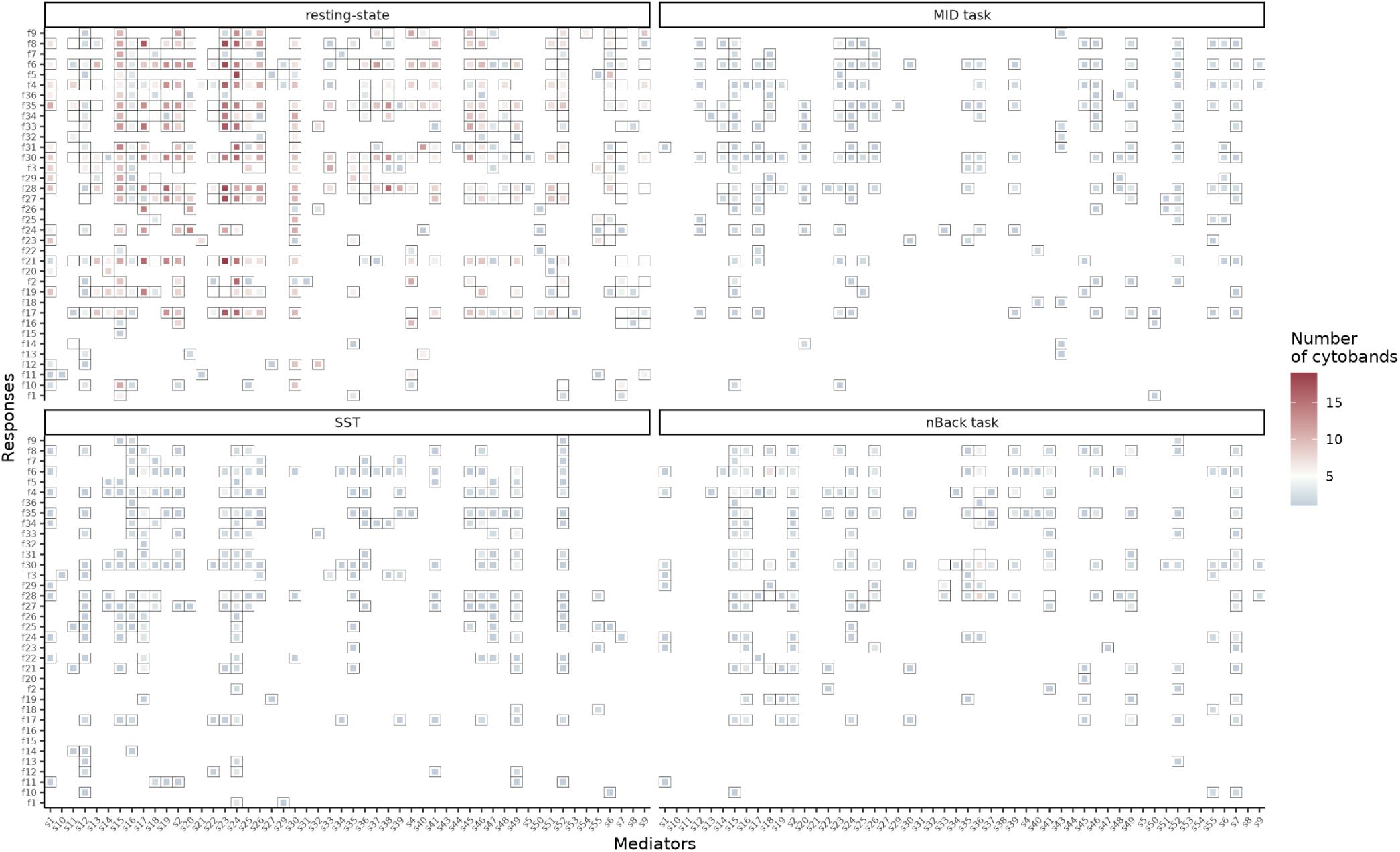
Number of cytobands associated with each combination of functional (y-axis) and structural (x-axis) network traits. The color gradient represents the number of cytobands, where deeper shades indicate a greater number of identified cytobands for that specific combination.

**Figure 5:**
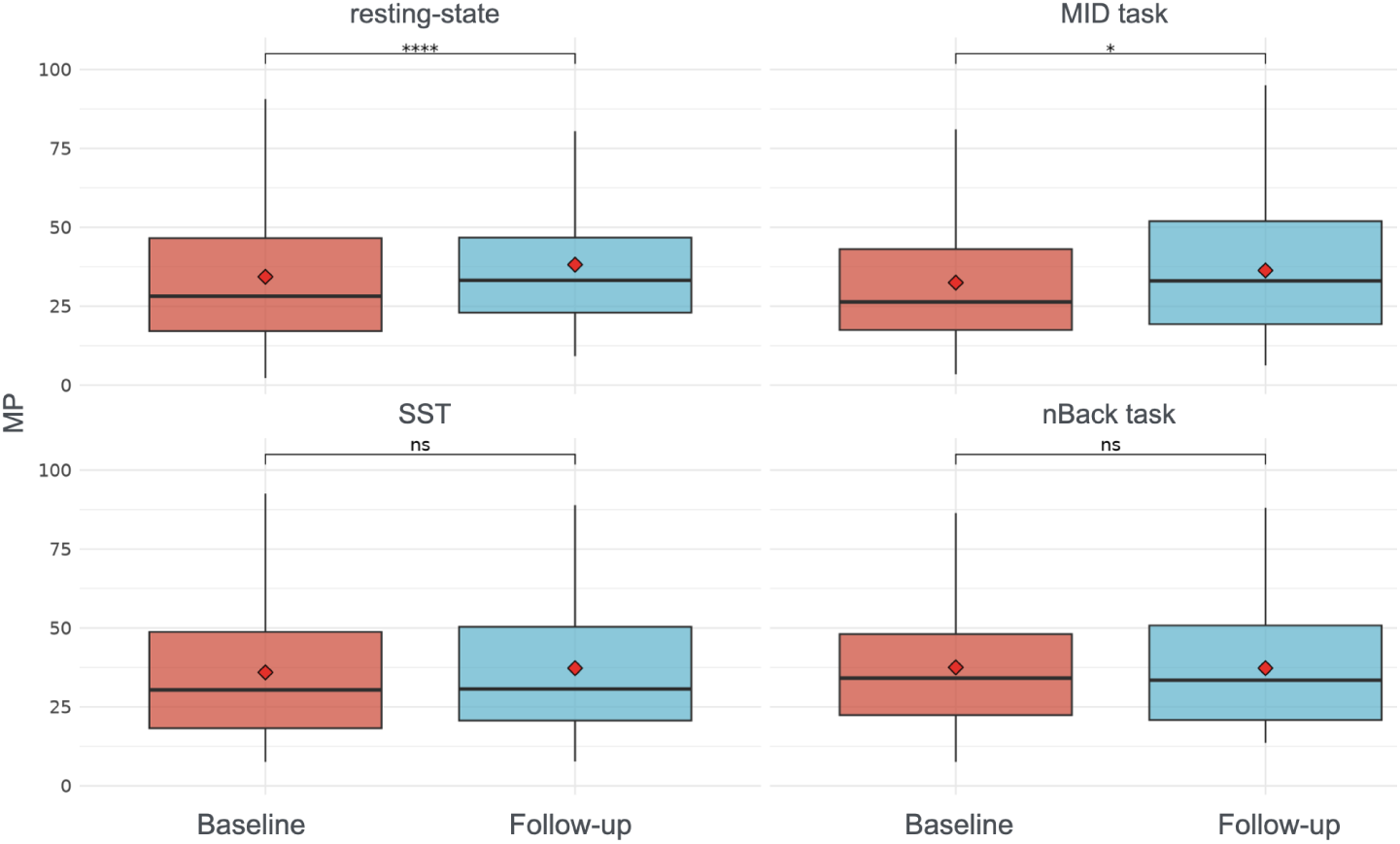
Boxplots of MP across the four cognitive states and two datasets at baseline and two-year follow-up. Red diamonds represent the mean MP values for each condition. The statistical significance of the Mann-Whitney U test is displayed at the top of each subplot, highlighting differences in MP distributions between the two datasets. Here “ns” indicates the mean difference in MP between the baseline data set and two-year follow-up dataset is not significant, “****” indicates p-value *<* 1e-04, and “*” indicates 0.01 *<* p-value *<* 0.05.

#### 3.2.2 Right frontal lobe-Mediated genetic effects on network-based FC

Previous studies have shown that certain genetic variants are associated with variations in cortical thickness and white matter microstructure (A. A. Joshi et al., 2011; Kochunov et al., 2011). These structural modifications may disrupt neural pathways and network communication, thereby affecting network-based FC. To better understand the causal mediation of SNPs, we selected a subset of SNPs from the independent lead SNP list or within LD blocks for each state and focused on significant mediation pathways involving the right frontal lobe, given that the right frontal lobe plays a pivotal role in mediating genetic effects on various neural networks, influencing a range of cognitive and motor functions. Below, we highlight our findings as presented in Table 2 along with relevant literature.

**Table 2:** Direct and indirect effects (×10^−3^) with 95% confidence intervals of SNP effects on functional neuroimaging traits, mediated by the right frontal lobe during resting state, MID task, SST task and nBack task. Abbreviations: DMN: Default mode network; SC-Cer: Subcortical-cerebellum.

| State | SNP | Outcome | Direct Effect (95% CI) | Indirect Effect (95% CI) |
| --- | --- | --- | --- | --- |
| Resting | rs73988141 | (DMN,DMN) | 0.84 (-16.30, 16.40) | -2.38 (-4.10, -1.07) |
|  |  | (Motor,Motor) | 1.11 (-2.85, 5.49) | -0.63 (-1.12, -0.22) |
|  |  | (SC-Cer, SC-Cer) | 2.15 (-2.55, 7.29) | -0.55 (-1.01, -0.17) |
|  |  | (Visual II,Visual II) | 7.70 (-17.90, 32.40) | -3.18 (-6.00, -1.00) |
|  |  | (Visual association, Visual association) | 6.85 (-4.59, 18.30) | -1.87 (-3.54, -0.70) |
| MID | rs73988140 | (SC-Cer SC-Cer) | -0.70 (-4.14, 2.73) | -0.66 (-1.15, -0.19) |
|  |  | (Visual association, Visual association) | -8.55 (-21.00, 3.87) | -1.66 (-3.17, -0.43) |
|  |  | (SC-Cer, Visual I) | -6.22 (-10.6, -1.85) | -1.10 (-1.99, -0.42) |
|  |  | (SC-Cer, Visual II) | -9.16 (-13.90, -4.28) | -0.78 (-1.52, -0.22) |
|  |  | (SC-Cer, Visual association) | -3.62 (-9.11, 2.04) | -1.27 (-2.19, -0.42) |
| SST | rs73988141 | (DMN,Visual II) | 7.75 (-5.20, 20.20) | -2.20 (-4.30, -0.62) |
| nBack | rs73988140 | (Visual I,Visual I) | -7.32 (-21.50, 6.68) | -1.78 (-3.82, -0.48) |
|  |  | (SC-Cer, Visual association) | -0.92 (-6.74, 5.11) | -0.91 (-1.86, -0.21) |

Firstly, in both the resting-state and MID tasks, we observed the right frontal lobe mediates genetic effects on the Subcortical-cerebellum (SC-Cer) network. This mediation is facilitated through cerebello-thalamo-cortical circuits, which integrate cognitive, motor, and emotional processes. The dorsolateral prefrontal cortex and orbitofrontal cortex interact with the thalamus, basal ganglia, and cerebellum, supporting executive control, working memory, and motor coordination (Middleton & Strick, 2000). fMRI studies show that the cerebellum contributes to intrinsic connectivity networks, particularly the central executive network, which involves the prefrontal cortex (Buckner et al., 2011).

Second, the right frontal lobe was observed to transmit indirect genetic effects to visual-related networks in the resting-state, MID task, and nBack task. This region is critically involved in higher-order visual processing, attention, and visuospatial functions, particularly within the dorsal and ventral visual pathways, which process spatial awareness and object recognition, respectively. The right frontal eye field, located in the dorsolateral prefrontal cortex, plays a central role in voluntary eye movements and attentional control over visual stimuli (Paus, 1996). Additionally, the right prefrontal cortex, including the inferior and middle frontal gyri, modulates visual perception through top-down cognitive control, influencing visual attention and working memory (Corbetta & Shulman, 2002).

The right frontal lobe is a key component of the Default mode network (DMN), associated with self-referential thinking, mind-wandering, and introspection. Within the DMN, the right medial prefrontal cortex plays a central role in autobiographical memory, social cognition, and decision-making (Raichle et al., 2001). FC studies indicate that the right frontal lobe interacts with the posterior cingulate cortex and precuneus, forming a core part of the DMN’s functional architecture (Greicius et al., 2003).

In addition, regarding motor functions, we observed the right frontal lobe mediated the pathway from genetics to intra-Motor network in resting-state. This result is consistent with existing findings that the right frontal lobe is involved in motor planning, execution, and control through interactions with cortical and subcortical structures. The primary motor cortex, located in the right precentral gyrus, is responsible for voluntary movements of the left side of the body, following the principle of contralateral motor control (Penfield & Boldrey, 1937). The right premotor cortex and supplementary motor area contribute to motor preparation, sequencing, and coordination, integrating sensory feedback to refine movement execution (Hikosaka et al., 2002). These regions are heavily interconnected with the basal ganglia, thalamus, and cerebellum, forming corticospinal and cortico-subcortical circuits that regulate movement precision and adaptation (Kelly & Strick, 2003).

In summary, the genetic effect could be transmitted to inter-network FC through the right frontal lobe. As discussed previously, the right frontal lobe influences several functional networks, including the DMN, Motor, Visual I, Visual II, Visual association, and SC-Cer networks. This raises the question of whether the right frontal lobe also influences inter-network functional traits. To address this, coupling between the right frontal lobe and various functional inter-network traits was examined to explore the relationship between structural and functional traits. For the MID task, the couplings between the right frontal lobe and (SC-Cer, Visual I) and (SC-Cer, Visual Association) were -0.14 and -0.15, respectively. In the SST, the coupling between the right frontal lobe and (DMN, Visual II) was -0.12, while for the nBack task, the coupling between the right frontal lobe and (SC-Cer, Visual Association) was -0.11. These results demonstrate relatively strong associations between these structural-functional network trait pairs, indicating that the right frontal lobe plays a significant role as a structural mediator in shaping functional connectivity across multiple networks. This suggests that its influence extends beyond isolated networks, contributing to broader network-level interactions that support complex brain functions.

### 3.3 Functional annotation and shared loci for causal genetic factors

#### 3.3.1 Gene mapping of mediation pathway SNPs

For the resting-state, after mapping the SNPs involved in significant NIE to their corresponding genes, we identified numerous genes that may influence brain development and neurodevelopmental disorders, including CREB1, DPM1, ADNP, and GABRG3, among others. CREB1 is involved in neuronal plasticity, learning, and memory, and its dysregulation has been linked to intellectual disabilities and cognitive impairments (Saura & Cardinaux, 2017). DPM1 is essential for glycosylation processes in the brain, and mutations in this gene lead to severe neurological deficits, developmental delays, and epilepsy (Barone et al., 2012). ADNP plays a key role in synaptic function and neurodevelopment, with mutations causing ADNP syndrome, which is characterized by intellectual disabilities, autism spectrum disorder, and motor impairments (Helsmoortel et al., 2014). GABRG3 is involved in inhibitory neurotransmission, and its alterations have been associated with autism spectrum disorder and epilepsy (C.-H. Chen et al., 2014).

Additionally, in order to explore the genetic effects on the functional networks through the structural mediators under the MID task, SNPs were mapped to their nearest genes, identifying a total of nine genes. Some of these genes, such as ROBO1, DAB1, and KCNK3, play crucial roles in neural processes. ROBO1 is essential for axon guidance, and its disruption has been linked to developmental dyslexia (Hannula-Jouppi et al., 2005). DAB1 is critical for neuronal migration, with alterations in its function associated with abnormal cortical development and neurodevelopmental disorders (Trommsdorff et al., 1999). KCNK3 contributes to the regulation of neuronal excitability, and its dysregulation has been implicated in cerebral ischemia (Muhammad et al., 2010).

Similarly, we identified several genes associated with the SST. Some candidate genes, such as ROBO1 and GRID1, are linked to brain function and neurological disorders. ROBO1 has been associated with developmental dyslexia, with genetic variations influencing reading ability (Hannula-Jouppi et al., 2005). It has also been studied in relation to brain lateralization and auditory processing (Lamminmäki et al., 2012). GRID1 has been implicated in schizophrenia, with altered expression patterns observed in individuals with the disorder (Fallin et al., 2005).

Furthermore, evidence suggests that several mapped genes associated with the nBack task are linked to neurodevelopmental and psychiatric disorders. For instance, variations in SP4 have been linked to schizophrenia and bipolar disorder, with studies indicating altered expression in the prefrontal cortex of affected individuals (Zhou et al., 2009). Additionally, structural brain abnormalities associated with DPYSL5 mutations suggest its involvement in brain connectivity and cognitive function (Desprez et al., 2023).

#### 3.3.2 EQTL profiling of causal SNPs

To distinguish SNPs that causally regulate gene expression from those that are merely associated due to linkage disequilibrium, eQTL profiling was conducted on the SNPs identified in the mediation analysis. Several SNPs were identified as eQTLs across different functional states, mapping them to genes implicated in brain function and neurodevelopment. The cis-eQTL results are displayed in Table 3.

**Table 3:** cis-eQTL results for selected significant lead SNPs obtained from GTEx/UKBEC brain database.

| State | SNP | Cytoband | p-value | Regulated genes |
| --- | --- | --- | --- | --- |
| resting-state | rs144067396 | p11.32 | 2.71E-05 | SMCHD1 |
|  | rs74101672 | q31.1 | 2.57E-05 | SLITRK6 |
|  | rs143925313 | q13.13 | 2.08E-05 | ADNP |
|  | rs143925313 | q13.13 | 1.27E-06 | DPM1 |
| MID task | rs1407504 | q22.33 | 1.33E-05 | COL15A1 |
| SST | rs58977077 | q31.3 | 4.24E-23 | FAM160A1 |
|  | rs58977077 | q31.3 | 5.34E-22 | PET112 |
|  | rs58977077 | q31.3 | 1.31E-15 | SH3D19 |
| nBack task | rs141217457 | q15.3 | 1.68E-05 | CATSPER2 |
|  | rs6060311 | q11.22 | 1.18E-04 | CEP250 |
|  | rs6060311 | q11.22 | 3.19E-05 | NCOA6 |

For the resting-state condition, rs143925313 (cytobands p11.32 and q13.13) and rs74101672 (q31.1), were associated with ADNP, DPM1, SMCHD1, and SLITRK6 in brain tissues (GTEx v8). ADNP and DPM1 (Section 3.3.1) are critical for synaptic function and glycosylation, while SLITRK6 modulates neuronal outgrowth and synaptic plasticity, with enriched expression in sensory and cognitive brain regions (Liu et al., 2020). A similar procedure was applied to the SNPs under the MID task. rs1407504 (q22.33) was mapped to COL15A1, encoding collagen XV, which localizes to brain endothelial cells and peripheral nerves. Collagen XV supports nerve maturation and C-fiber assembly, processes essential for sensory signaling (Rasi et al., 2010). In the SST, the leading SNP rs58977077 (q31.3) was associated with SH3D19, PET112, and FAM160A1. SH3D19 has been tied to Alzheimer’s disease via homozygosity analyses (Ghani et al., 2015), PET112 to PTSD in transcriptome-wide studies (Wingo et al., 2022), and FAM160A1 to cerebrovascular integrity in subarachnoid hemorrhage (Yamada et al., 2018).

Finally, the nBack task implicated rs141217457 (q15.3) and rs6060311 (q11.22) as eQTLs for NCOA6, CEP250, and CATSPER2. While CEP250 dysfunction disrupts centrosome cohesion and is tied to neurodevelopmental disorders (Floriot et al., 2015), the roles of NCOA6 (a transcriptional coactivator) and CATSPER2 (a sperm-specific ion channel) in cognition remain uncharacterized.

A complete list of mediation pathways involving eQTLs is provided in Supplemental Tables S3–S6. Figure 6 visualizes the mediation pathways for selected lead SNPs in each state. This visualization demonstrated the indirect influence of each SNP on FC network traits, mediated through SC network traits, providing insights into the genetic mechanisms underlying brain function.

**Figure 6:**
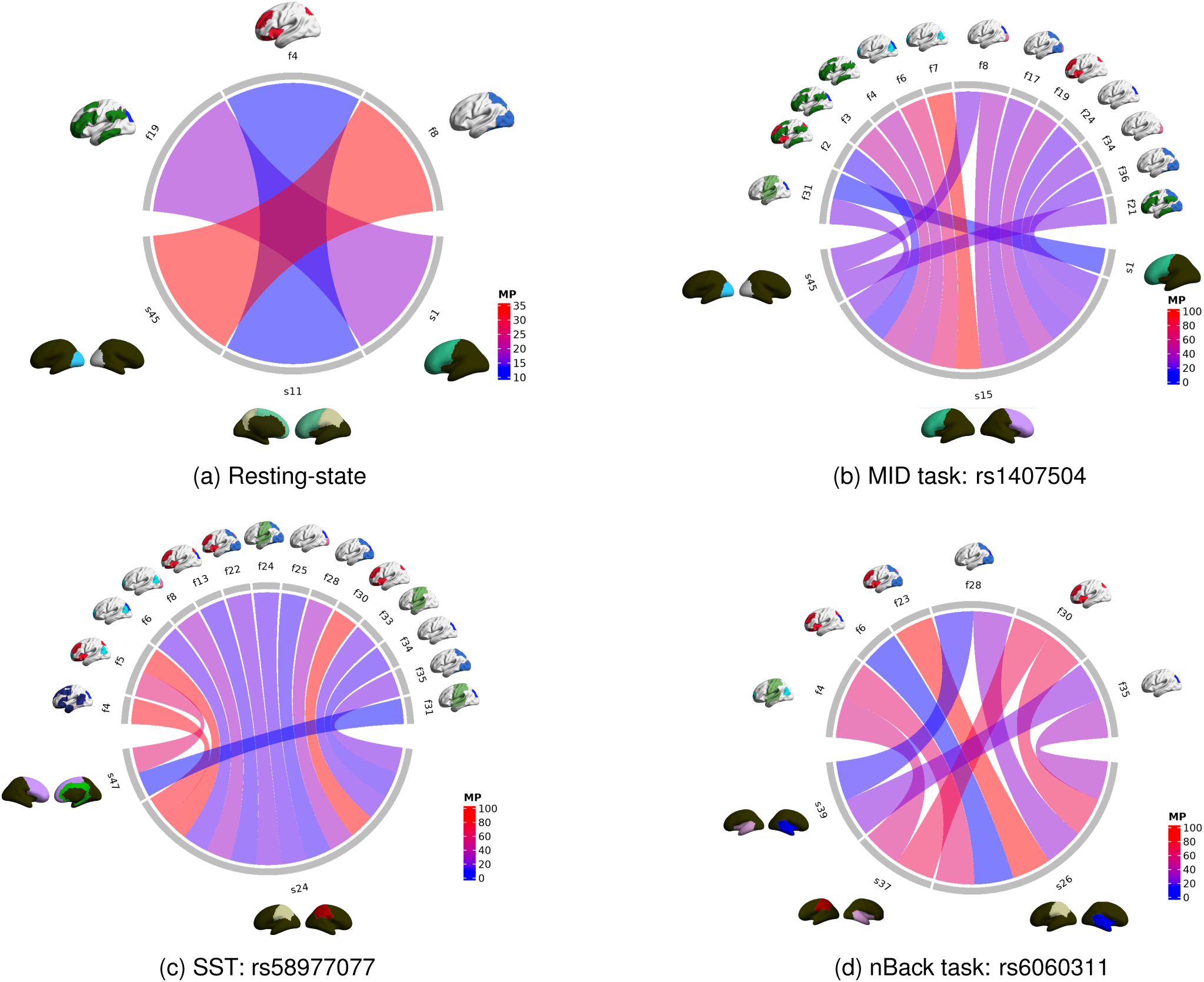
Selected lead SNP(s) for each cognitive state, with edge colors representing the proportion of the intermediate effect.

### 3.4 Evaluation of mediation consistency on two-year follow-up data

#### 3.4.1 Static genetic mediation pathways across time

To assess the robustness and reproducibility of our findings and prioritize stable genetic effects on SC and FC network traits, we conducted the same analyses using the two-year follow-up data, with a focus on identifying genetic influences that are stable and less susceptible to neurodevelopmental fluctuations. In addition to the GWAS replication analyses in Section 3.1.1, we conducted the replication analyses of significant mediator-to-response (SC mediator → FC network trait) pathways among the significant NIEs, providing further insight into the temporal stability of mediation effects over time.

Our findings indicated that the resting-state condition exhibited the highest replication rate, highlighting the relative stability of intrinsic brain network mediation effects. Specifically, we identified 526 significant SC mediator → FC network trait pathways in the baseline set, involving 51 SC and 35 FC network traits. In the replication analyses, 164 significant pathways were detected, with 110 overlapping with the baseline set. Notably, 34 out of 35 SC network traits and 28 out of 29 FC network traits in the replication analyses were also present in the baseline set. This substantial level of replication in mediation pathways highlighted the capability of our mediation analysis framework to uncover fundamental and stable neurogenetic mechanisms.

The MID, SST, and nBack tasks also demonstrated varying levels of replication. For the MID task, 227 significant pathways were identified in the baseline dataset, involving 34 SC and 32 FC network traits. The replication analyses revealed 70 pathways, with 32 overlapping with the baseline set. Among the traits, 22 SC and 17 FC network traits were consistently replicated. For the SST, 273 pathways were detected in the baseline dataset, including 39 SC and 33 FC network traits. The replication analyses identified 64 pathways, with 25 replicating the baseline set. Most network traits—22 SC and 21 FC—were retained across both datasets. For the nBack task, 221 pathways were found in the baseline dataset, consisting of 39 SC and 33 FC network traits. The replication analyses identified 39 pathways, 12 of which were replicated in the baseline set. While only 10 SC and 15 FC network traits were retained across datasets, the lower replication rate may reflect dynamic changes in task-related connectivity over time.

Overall, these results indicated moderate to high levels of replication across functional states, with the resting-state condition showing the highest consistency, underscoring the relative stability of intrinsic brain network mediation effects.

#### 3.4.2 Temporal refinement of neuroimaging traits during neurodevelopment

Our mediation analysis revealed moderate to high replication rates of SC-mediated pathways linking SNPs and FC network traits across the baseline and two-year follow-up datasets. Nonetheless, adolescence is a period marked by dynamic neurodevelopment, including synaptic pruning, myelination, and shifts in network organization (Fair et al., 2009; Simmonds et al., 2014). Genetic effects on brain connectivity can be modulated by age, pubertal status, and hormonal changes, potentially contributing to discrepancies between the baseline and two-year follow-up datasets (Simmonds et al., 2014; Tamnes et al., 2017; B. Zhao et al., 2021). While our primary aim was to detect stable genetic associations, acknowledging these dynamic processes is important, as they may influence the strength or presence of genetic mediation pathways over time.

To explore temporal variations, we applied linear mixed-effects models to the neuroimaging traits, incorporating random subject effects. Figure 7 presents volcano plots illustrating significant changes in SC and FC network traits between these two time points. Corresponding boxplots reveal that some traits exhibited developmental shifts, consistent with the neuroplastic changes characteristic of this age range. Despite these variations, the replicated SC-mediated genetic pathways to FC networks suggested that certain genetic influences on brain network architecture are robust to neurodevelopmental transitions.

**Figure 7:**
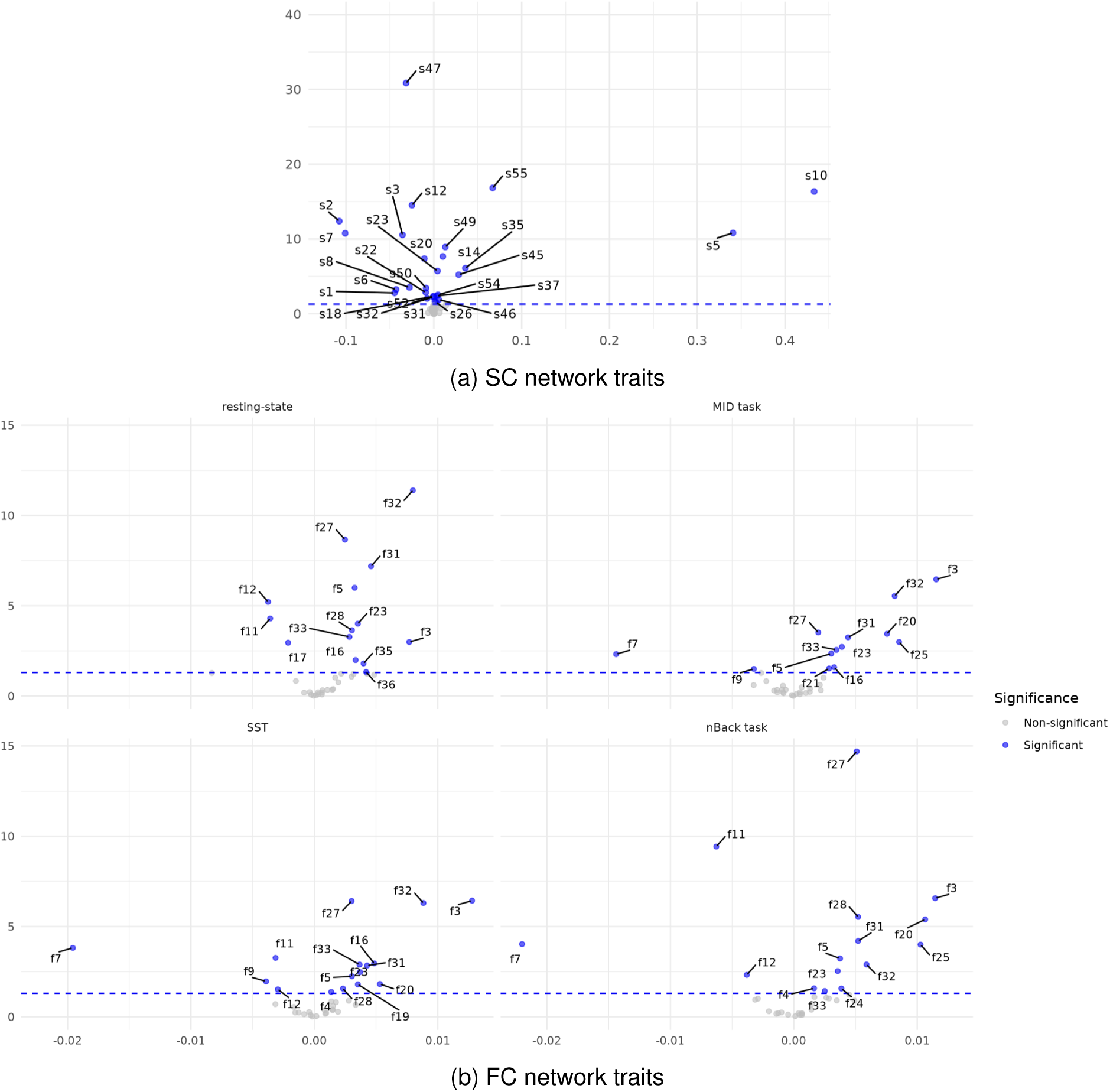
Volcano plots of SC and FC network traits across the four functional states for baseline and two-year follow-up data. The color represents the statistical significance of the time effect, indicating differences in the derived network traits between these two time points.

## 4 Discussion

In this study, we conducted SNP-to-multi-type brain network mediation analyses to identify causal genetic variants that influence brain functional network architectures by modulating structural connectivity networks. By first establishing genetic associations with SC and FC network traits, we employed causal mediation modeling to uncover genetic pathways that impose direct effects on functional networks and indirect effects mediated through structural networks. By incorporating a comprehensive set of SC and FC traits across the entire brain, our analysis enabled the identification of remote coupling effects and potential effect rerouting from structural networks to functional connectivity at different brain areas shaped by genetic factors. Furthermore, through genetic functional annotation and eQTL analyses, we demonstrated the role of causal genetic variants in brain development and their potential contribution to neurodegenerative disease susceptibility. Replication analyses at a two-year follow-up confirmed the widespread and stable influence of genetic factors on SC and FC network alterations during neurodevelopment, while also highlighting the dynamic refinement of both genetic contributions and imaging genetic effect pathways over time.

Our work presented several exciting opportunities for further study. First, in this work, we employed the Shen 268-node atlas for FC in both resting-state fMRI and task-based fMRI. However, using the same atlas for all modalities presents a potential challenge, as resting-state networks may not fully capture the spatial specificity of task-evoked activations. This mismatch between functional parcellations and task-driven neural responses could lead to inaccuracies in the interpretation of task-related connectivity patterns. The Shen 268-node parcellation is optimized for intrinsic connectivity patterns but may inadvertently group subregions that exhibit distinct activation profiles during tasks, reducing sensitivity to localized functional differences. Furthermore, task-related activations and deactivations—such as those observed in the Default mode network—may not be adequately represented within a framework derived from resting-state connectivity, increasing the risk of misinterpreting task effects. Additionally, network boundaries established through resting-state connectivity may not generalize well across diverse cognitive tasks, limiting their applicability for task-specific analyses.

Second, this paper utilized two-year follow-up data for replication analyses to identify stable and dynamic neurogenetic mechanisms. However, extending our mediation framework to fully account for the longitudinal nature of the data would be valuable. Since both mediators (SC network traits) and outcomes (FC network traits) are repeatedly measured, lagged mediation effects may carry over to the next time point. This could be potentially addressed through growth curve mediation models, allowing for a more nuanced understanding of how genetic influences shape brain network development over time.

Finally, while neuroimaging genetics helps identify intermediate traits linking genetic variation to behavior, integrating genetic, neuroimaging, and behavioral data is essential for establishing causal relationships. A multimodal approach would offer deeper insights into how genetic variants shape brain function and behavior. Similarly, applying our framework to diverse datasets like the Human Connectome Project and UK Biobank could reveal population-specific and shared neurogenetic mechanisms, enhancing our understanding of SC-mediated genetic influences on FC across ancestries.

## Data and Code Availability

The original imaging, genomics and behavioral data are from Adolescent Brain Cognitive Development study through NIMH Data Archive (https://nda.nih.gov/abcd) upon approvals.

## Author Contributions

S.G. and Y.Z. designed the study. S.G., S.D., X.Z. and Z.G. processed, analyzed and visualized the data. Y.Z. acquired the funding support. All authors contributed to write and edit the paper.

## Funding

This work was partially supported by the National Institutes of Health under awards RF1AG081413, R01EB034720 and R01AG068191.

## Declaration of Competing Interests

The authors declare no competing interests.

## Data Availability

The data are stored in the NIMH Data Archive (NDA;https://nda.nih.gov/abcd).

## Acknowledgements

Data used in the preparation of this article were obtained from the Adolescent Brain Cognitive DevelopmentSM (ABCD) Study (https://abcdstudy.org), held in the NIMH Data Archive (NDA). This is a multisite, longitudinal study designed to recruit more than 10,000 children age 9-10 and follow them over 10 years into early adulthood. The ABCD Study ® is supported by the National Institutes of Health and additional federal partners under award numbers U01DA041048, U01DA050989, U01DA051016, U01DA041022, U01DA051018, U01DA051037, U01DA050987, U01DA041174, U01DA041106, U01DA041117, U01DA041028, U01DA041134, U01DA050988, U01DA051039, U01DA041156, U01DA041025, U01DA041120, U01DA051038, U01DA041148, U01DA041093, U01DA041089, U24DA041123, U24DA041147. A full list of supporters is available at https://abcdstudy.org/federal-partners.html. A listing of participating sites and a complete listing of the study investigators can be found at https://abcdstudy.org/consortium_members/. ABCD consortium investigators designed and implemented the study and/or provided data but did not necessarily participate in the analysis or writing of this report. This manuscript reflects the views of the authors and may not reflect the opinions or views of the NIH or ABCD consortium investigators. The ABCD data repository grows and changes over time. The ABCD data used in this report came from release 5.1. DOIs can be found at http://dx.doi.org/10.15154/z563-zd24.

## Notes

### Competing Interest Statement

The authors have declared no competing interest.

### Author Declarations

The Institutional Review Board of the University of California, San Diego gave ethical approval for the Adolescent Brain Cognitive Development (ABCD) Study. Use of this dataset was approved through data use certification from the NIMH Data Archive (NDA).

## References

Alwin, D. F., & Hauser, R. M. (1975). The decomposition of effects in path analysis. American Sociological Review, 37–47.

Barone, R., Aiello, C., Race, V., Morava, E., Foulquier, F., Riemersma, M., Passarelli, C., Concolino, D., Carella, M., Santorelli, F., et al. (2012). Dpm2-cdg: A muscular dystrophy–dystroglycanopathy syndrome with severe epilepsy. Annals of Neurology, 72(4), 550–558.

Bi, X., Yang, L., Li, T., Wang, B., Zhu, H., & Zhang, H. (2017). Genome-wide mediation analysis of psychiatric and cognitive traits through imaging phenotypes. Human Brain Mapping, 38(8), 4088–4097.

Bienvenu, O. J., Davydow, D. S., & Kendler, K. S. (2011). Psychiatric ‘diseases’ versus behavioral disorders and degree of genetic influence. Psychological Medicine, 41(1), 33–40.

Brouwer, R. M., Klein, M., Grasby, K. L., Schnack, H. G., Jahanshad, N., Teeuw, J., Thomopoulos, S. I., Sprooten, E., Franz, C. E., Gogtay, N., et al. (2022). Genetic variants associated with longitudinal changes in brain structure across the lifespan. Nature Neuroscience, 25(4), 421–432.

Buckner, R. L., Krienen, F. M., Castellanos, A., Diaz, J. C., & Yeo, B. T. (2011). The organization of the human cerebellum estimated by intrinsic functional connectivity. Journal of Neurophysiology, 106(5), 2322– 2345.

Bullmore, E., & Sporns, O. (2009). Complex brain networks: Graph theoretical analysis of structural and functional systems. Nature Reviews Neuroscience, 10(3), 186–198.

Cao, R., Wang, X., Gao, Y., Li, T., Zhang, H., Hussain, W., Xie, Y., Wang, J., Wang, B., & Xiang, J. (2020). Abnormal anatomical rich-club organization and structural–functional coupling in mild cognitive impairment and alzheimer’s disease. Frontiers in Neurology, 11, 53.

Casey, B., Cannonier, T., Conley, M. I., Cohen, A. O., Barch, D. M., Heitzeg, M. M., Soules, M. E., Teslovich, T., Dellarco, D. V., Garavan, H., Orr, C. A., Wager, T. D., Banich, M. T., Speer, N. K., Sutherland, M. T., Riedel, M. C., Dick, A. S., Bjork, J. M., Thomas, K. M., . . . Dale, A. M. (2018). The Adolescent Brain Cognitive Development (ABCD) study: Imaging acquisition across 21 sites [The Adolescent Brain Cognitive Development (ABCD) Consortium: Rationale, Aims, and Assessment Strategy]. Developmental Cognitive Neuroscience, 32, 43–54.

Chen, C.-H., Huang, C.-C., Cheng, M.-C., Chiu, Y.-N., Tsai, W.-C., Wu, Y.-Y., Liu, S.-K., & Gau, S. S.-F. (2014). Genetic analysis of gabrb3 as a candidate gene of autism spectrum disorders. Molecular Autism, 5, 1–13.

Chen, T., Mandal, A., Zhu, H., & Liu, R. (2022). Imaging genetic based mediation analysis for human cognition. Frontiers in Neuroscience, 16, 824069.

Chen, Z., Wu, X., Yang, Q., Zhao, H., Ying, H., Liu, H., Wang, C., Zheng, R., Lin, H., Wang, S., et al. (2024). The effect of sglt2 inhibition on brain-related phenotypes and aging: A drug target mendelian randomization study. The Journal of Clinical Endocrinology & Metabolism, dgae635.

Cohen, A., Conley, M., Dellarco, D., & Casey, B. (2016). The impact of emotional cues on short-term and long-term memory during adolescence. *Proceedings of the Society for Neuroscience. San Diego, CA. November*.

Cole, M., Murray, K., St-Onge, E., Risk, B., Zhong, J., Schifitto, G., Descoteaux, M., & Zhang, Z. (2021). Surface-based connectivity integration: An atlas-free approach to jointly study functional and structural connectivity. Human Brain Mapping, 42(11), 3481–3499.

Consortium, G., Ardlie, K. G., Deluca, D. S., Segrè, A. V., Sullivan, T. J., Young, T. R., Gelfand, E. T., Trowbridge, C. A., Maller, J. B., Tukiainen, T., et al. (2015). The genotype-tissue expression (gtex) pilot analysis: Multitissue gene regulation in humans. Science, 348(6235), 648–660.

Corbetta, M., & Shulman, G. L. (2002). Control of goal-directed and stimulus-driven attention in the brain. Nature Reviews Neuroscience, 3(3), 201–215.

Coutts, J. J., & Hayes, A. F. (2023). Questions of value, questions of magnitude: An exploration and application of methods for comparing indirect effects in multiple mediator models. Behavior Research Methods, 55(7), 3772–3785.

Dai, W., Zhang, Z., Song, P., Zhang, H., & Zhao, Y. (2024). Heritability and genetic contribution analysis of structural-functional coupling in human brain. Imaging Neuroscience, 2, 1–19.

Davies, G., Tenesa, A., Payton, A., Yang, J., Harris, S. E., Liewald, D., Ke, X., Le Hellard, S., Christoforou, A., Luciano, M., et al. (2011). Genome-wide association studies establish that human intelligence is highly heritable and polygenic. Molecular Psychiatry, 16(10), 996–1005.

Desprez, F., Ung, D. C., Vourc’h, P., Jeanne, M., & Laumonnier, F. (2023). Contribution of the dihydropyrimidinase-like proteins family in synaptic physiology and in neurodevelopmental disorders. Frontiers in Neuroscience, 17, 1154446.

Fair, D. A., Cohen, A. L., Power, J. D., Dosenbach, N. U., Church, J. A., Miezin, F. M., Schlaggar, B. L., & Petersen, S. E. (2009). Functional brain networks develop from a “local to distributed” organization. PLoS Computational Biology, 5(5), e1000381.

Fallin, M. D., Lasseter, V. K., Avramopoulos, D., Nicodemus, K. K., Wolyniec, P. S., McGrath, J. A., Steel, G., Nestadt, G., Liang, K.-Y., Huganir, R. L., et al. (2005). Bipolar i disorder and schizophrenia: A 440– single-nucleotide polymorphism screen of 64 candidate genes among ashkenazi jewish case-parent trios. The American Journal of Human Genetics, 77 (6), 918–936.

Floriot, S., Vesque, C., Rodriguez, S., Bourgain-Guglielmetti, F., Karaiskou, A., Gautier, M., Duchesne, A., Barbey, S., Fritz, S., Vasilescu, A., et al. (2015). C-nap1 mutation affects centriole cohesion and is associated with a seckel-like syndrome in cattle. Nature Communications, 6(1), 6894.

Friston, K. J. (1994). Functional and effective connectivity in neuroimaging: A synthesis. Human Brain Mapping, 2(1-2), 56–78.

Garavan, H., Bartsch, H., Conway, K., Decastro, A., Goldstein, R., Heeringa, S., Jernigan, T., Potter, A., Thompson, W., & Zahs, D. (2018). Recruiting the abcd sample: Design considerations and procedures. Developmental Cognitive Neuroscience, 32, 16–22.

Ghani, M., Reitz, C., Cheng, R., Vardarajan, B. N., Jun, G., Sato, C., Naj, A., Rajbhandary, R., Wang, L.-S., Valladares, O., et al. (2015). Association of long runs of homozygosity with alzheimer disease among african american individuals. JAMA Neurology, 72(11), 1313–1323.

Glasser, M. F., Sotiropoulos, S. N., Wilson, J. A., Coalson, T. S., Fischl, B., Andersson, J. L., Xu, J., Jbabdi, S., Webster, M., Polimeni, J. R., et al. (2013). The minimal preprocessing pipelines for the human connectome project. Neuroimage, 80, 105–124.

Goldberg, T. E., & Weinberger, D. R. (2004). Genes and the parsing of cognitive processes. Trends in Cognitive Sciences, 8(7), 325–335.

Greicius, M. D., Krasnow, B., Reiss, A. L., & Menon, V. (2003). Functional connectivity in the resting brain: A network analysis of the default mode hypothesis. Proceedings of the National Academy of Sciences, 100(1), 253–258.

Gu, Z., Jamison, K. W., Sabuncu, M. R., & Kuceyeski, A. (2021). Heritability and interindividual variability of regional structure-function coupling. Nature Communications, 12(1), 4894.

Hannula-Jouppi, K., Kaminen-Ahola, N., Taipale, M., Eklund, R., Nopola-Hemmi, J., Kääriäinen, H., & Kere, J. (2005). The axon guidance receptor gene ROBO1 is a candidate gene for developmental dyslexia. PLoS Genetics, 1(4), e50.

Helsmoortel, C., Vulto-van Silfhout, A. T., Coe, B. P., Vandeweyer, G., Rooms, L., van den Ende, J., Schuurs-Hoeijmakers, J. H., Marcelis, C. L., Willemsen, M. H., Vissers, L. E., et al. (2014). A swi/snf-related autism syndrome caused by de novo mutations in adnp. Nature Genetics, 46(4), 380–384.

Hikosaka, O., Nakamura, K., Sakai, K., & Nakahara, H. (2002). Central mechanisms of motor skill learning. Current Opinion in Neurobiology, 12(2), 217–222.

Hurles, M. E., Dermitzakis, E. T., & Tyler-Smith, C. (2008). The functional impact of structural variation in humans. Trends in Genetics, 24(5), 238–245.

Joshi, A., Scheinost, D., Okuda, H., Belhachemi, D., Murphy, I., Staib, L. H., & Papademetris, X. (2011). Unified framework for development, deployment and robust testing of neuroimaging algorithms. Neuroinformatics, 9, 69–84.

Joshi, A. A., Lepore, N., Joshi, S. H., Lee, A. D., Barysheva, M., Stein, J. L., McMahon, K. L., Johnson, K., de Zubicaray, G. I., Martin, N. G., et al. (2011). The contribution of genes to cortical thickness and volume. Neuroreport, 22(3), 101–105.

Jun, S., Altmann, A., & Sadaghiani, S. (2025). Modulatory neurotransmitter genotypes shape dynamic functional connectome reconfigurations. Journal of Neuroscience.

Kelly, R. M., & Strick, P. L. (2003). Cerebellar loops with motor cortex and prefrontal cortex of a nonhuman primate. Journal of Neuroscience, 23(23), 8432–8444.

Knutson, B., Westdorp, A., Kaiser, E., & Hommer, D. (2000). Fmri visualization of brain activity during a monetary incentive delay task. Neuroimage, 12(1), 20–27.

Kochunov, P., Glahn, D. C., Nichols, T. E., Winkler, A. M., Hong, E. L., Holcomb, H. H., Stein, J. L., Thompson, P. M., Curran, J. E., Carless, M. A., et al. (2011). Genetic analysis of cortical thickness and fractional anisotropy of water diffusion in the brain. Frontiers in Neuroscience, 5, 120.

Lam, M., Awasthi, S., Watson, H. J., Goldstein, J., Panagiotaropoulou, G., Trubetskoy, V., Karlsson, R., Frei, O., Fan, C.-C., De Witte, W., et al. (2020). Ricopili: Rapid imputation for consortias pipeline. Bioinformatics, 36(3), 930–933.

Lamminmäki, S., Massinen, S., Nopola-Hemmi, J., Kere, J., & Hari, R. (2012). Human ROBO1 regulates interaural interaction in auditory pathways. Journal of Neuroscience, 32(3), 966–971.

Lee, H., Chen, C., Kochunov, P., Hong, L. E., & Chen, S. (2024). A new multiple-mediator model maximally uncovering the mediation pathway: Evaluating the role of neuroimaging measures in age-related cognitive decline. The Annals of Applied Statistics, 18(4), 2775–2795.

Lindquist, M. A. (2012). Functional causal mediation analysis with an application to brain connectivity. Journal of the American Statistical Association, 107 (500), 1297–1309.

Liu, W., Zhang, X., Deng, Z., Li, G., Zhang, R., Yang, Z., Che, F., Liu, S., & Li, H. (2020). The role of slitrk6 in the pathogenesis of tourette syndrome: From the conclusion of a family-based study in the chinese han population. The Journal of Gene Medicine, 22(6), e3173.

Logan, G. D. (1994). Spatial attention and the apprehension of spatial relations. Journal of Experimental Psychology: Human Perception and Performance, 20(5), 1015.

Meyer-Lindenberg, A., & Weinberger, D. R. (2006). Intermediate phenotypes and genetic mechanisms of psychiatric disorders. Nature Reviews Neuroscience, 7 (10), 818–827.

Middleton, F. A., & Strick, P. L. (2000). Basal ganglia and cerebellar loops: Motor and cognitive circuits. Brain Research Reviews, 31(2-3), 236–250.

Mu, S., Rathore, S., Bao, J., Yang, S., Hooli, B., Shen, L., & ADNI. (2024). Genome-wide mediation analysis reveals amyloid and tau imaging genetics patterns in alzheimer’s disease. Alzheimer’s & Dementia, 20, e089710.

Muhammad, S., Aller, M. I., Maser-Gluth, C., Schwaninger, M., & Wisden, W. (2010). Expression of the kcnk3 potassium channel gene lessens the injury from cerebral ischemia, most likely by a general influence on blood pressure. Neuroscience, 167 (3), 758–764.

Park, H.-J., & Friston, K. (2013). Structural and functional brain networks: From connections to cognition. Science, 342(6158), 1238411.

Paus, T. (1996). Location and function of the human frontal eye-field: A selective review. Neuropsychologia, 34(6), 475–483.

Penfield, W., & Boldrey, E. (1937). Somatic motor and sensory representation in the cerebral cortex of man as studied by electrical stimulation. Brain, 60(4), 389–443.

Purcell, S., Neale, B., Todd-Brown, K., Thomas, L., Ferreira, M. A., Bender, D., Maller, J., Sklar, P., De Bakker, P. I., Daly, M. J., et al. (2007). Plink: A tool set for whole-genome association and population-based linkage analyses. The American Journal of Human Genetics, 81(3), 559–575.

Raichle, M. E., MacLeod, A. M., Snyder, A. Z., Powers, W. J., Gusnard, D. A., & Shulman, G. L. (2001). A default mode of brain function. Proceedings of the National Academy of Sciences, 98(2), 676–682.

Ramasamy, A., Trabzuni, D., Guelfi, S., Varghese, V., Smith, C., Walker, R., De, T., Coin, L. J. M., de Silva, R., Cookson, M. R., Singleton, A. B., Hardy, J., Ryten, M., & Weale, M. E. (2014). Genetic variability in the regulation of gene expression in ten regions of the human brain. Nature Neuroscience, 17, 1418– 1428.

Rasi, K., Hurskainen, M., Kallio, M., Stavén, S., Sormunen, R., Heape, A. M., Avila, R. L., Kirschner, D., Muona, A., Tolonen, U., et al. (2010). Lack of collagen xv impairs peripheral nerve maturation and, when combined with laminin-411 deficiency, leads to basement membrane abnormalities and sensorimotor dysfunction. Journal of Neuroscience, 30(43), 14490–14501.

Satterthwaite, T. D., Elliott, M. A., Gerraty, R. T., Ruparel, K., Loughead, J., Calkins, M. E., Eickhoff, S. B., Hakonarson, H., Gur, R. C., Gur, R. E., et al. (2013). An improved framework for confound regression and filtering for control of motion artifact in the preprocessing of resting-state functional connectivity data. Neuroimage, 64, 240–256.

Saura, C. A., & Cardinaux, J.-R. (2017). Emerging roles of creb-regulated transcription coactivators in brain physiology and pathology. Trends in Neurosciences, 40(12), 720–733.

Schneider, V. A., Graves-Lindsay, T., Howe, K., Bouk, N., Chen, H.-C., Kitts, P. A., Murphy, T. D., Pruitt, K. D., Thibaud-Nissen, F., Albracht, D., et al. (2017). Evaluation of grch38 and de novo haploid genome assemblies demonstrates the enduring quality of the reference assembly. Genome Research, 27 (5), 849–864.

Shen, L., & Thompson, P. M. (2019). Brain imaging genomics: Integrated analysis and machine learning. Proceedings of the IEEE, 108(1), 125–162.

Shen, X., Tokoglu, F., Papademetris, X., & Constable, R. T. (2013). Groupwise whole-brain parcellation from resting-state fmri data for network node identification. Neuroimage, 82, 403–415.

Simmonds, D. J., Hallquist, M. N., Asato, M., & Luna, B. (2014). Developmental stages and sex differences of white matter and behavioral development through adolescence: A longitudinal diffusion tensor imaging (dti) study. Neuroimage, 92, 356–368.

Stam, C. J. (2014). Modern network science of neurological disorders. Nature Reviews Neuroscience, 15(10), 683–695.

St-Onge, E., Daducci, A., Girard, G., & Descoteaux, M. (2018). Surface-enhanced tractography (set). NeuroImage, 169, 524–539.

Sun, Y., Dai, Z., Li, J., Collinson, S. L., & Sim, K. (2017). Modular-level alterations of structure–function coupling in schizophrenia connectome. Human Brain Mapping, 38(4), 2008–2025.

Tamnes, C. K., Herting, M. M., Goddings, A.-L., Meuwese, R., Blakemore, S.-J., Dahl, R. E., Güroğlu, B., Raznahan, A., Sowell, E. R., Crone, E. A., et al. (2017). Development of the cerebral cortex across adolescence: A multisample study of inter-related longitudinal changes in cortical volume, surface area, and thickness. Journal of Neuroscience, 37 (12), 3402–3412.

Teeuw, J., Mota, N. R., Klein, M., Blankenstein, N. E., Tielbeek, J. J., Jansen, L. M., Franke, B., & Pol, H. E. H. (2023). Polygenic risk scores and brain structures both contribute to externalizing behavior in childhood-a study in the adolescent brain and cognitive development (abcd) cohort. Neuroscience Applied, 2, 101128.

Tian, X., Li, F., Shen, L., Esserman, D., & Zhao, Y. (2024). Bayesian pathway analysis over brain network mediators for survival data. Biometrics, 80(4), ujae132.

Tissink, E., Werme, J., de Lange, S. C., Savage, J. E., Wei, Y., de Leeuw, C. A., Nagel, M., Posthuma, D., & van den Heuvel, M. P. (2023). The genetic architectures of functional and structural connectivity properties within cerebral resting-state networks. Eneuro, 10(4).

Trommsdorff, M., Gotthardt, M., Hiesberger, T., Shelton, J., Stockinger, W., Nimpf, J., Hammer, R. E., Richardson, J. A., & Herz, J. (1999). Reeler/disabled-like disruption of neuronal migration in knockout mice lacking the vldl receptor and apoe receptor 2. Cell, 97 (6), 689–701.

Van Essen, D. C. (2005). A population-average, landmark-and surface-based (pals) atlas of human cerebral cortex. Neuroimage, 28(3), 635–662.

VanderWeele, T., & Vansteelandt, S. (2009). Conceptual issues concerning mediation, interventions and composition. Statistics and its Interface, 2, 457–468.

Wang, S., Constable, T., Zhang, H., & Zhao, Y. (2024). Heterogeneity analysis on multi-state brain functional connectivity and adolescent neurocognition. Journal of the American Statistical Association, 119(546), 851–863.

Watanabe, K., Taskesen, E., Van Bochoven, A., & Posthuma, D. (2017). Functional mapping and annotation of genetic associations with fuma. Nature Communications, 8(1), 1826.

Wingo, T. S., Gerasimov, E. S., Liu, Y., Duong, D. M., Vattathil, S. M., Lori, A., Gockley, J., Breen, M. S., Maihofer, A. X., Nievergelt, C. M., et al. (2022). Integrating human brain proteomes with genome-wide association data implicates novel proteins in post-traumatic stress disorder. Molecular Psychiatry, 27 (7), 3075–3084.

Yamada, Y., Kato, K., Oguri, M., Horibe, H., Fujimaki, T., Yasukochi, Y., Takeuchi, I., & Sakuma, J. (2018). Identification of nine genes as novel susceptibility loci for early-onset ischemic stroke, intracerebral hemorrhage, or subarachnoid hemorrhage. Biomedical Reports, 9(1), 8–20.

Yang, Z., Zuo, X.-N., McMahon, K. L., Craddock, R. C., Kelly, C., de Zubicaray, G. I., Hickie, I., Bandettini, P. A., Castellanos, F. X., Milham, M. P., et al. (2016). Genetic and environmental contributions to functional connectivity architecture of the human brain. Cerebral Cortex, 26(5), 2341–2352.

Yeo, B. T., Krienen, F. M., Sepulcre, J., Sabuncu, M. R., Lashkari, D., Hollinshead, M., Roffman, J. L., Smoller, J. W., Zöllei, L., Polimeni, J. R., et al. (2011). The organization of the human cerebral cortex estimated by intrinsic functional connectivity. Journal of Neurophysiology.

Zhang, Z., Liao, W., Chen, H., Mantini, D., Ding, J.-R., Xu, Q., Wang, Z., Yuan, C., Chen, G., Jiao, Q., et al. (2011). Altered functional–structural coupling of large-scale brain networks in idiopathic generalized epilepsy. Brain, 134(10), 2912–2928.

Zhao, B., Li, T., Smith, S. M., Xiong, D., Wang, X., Yang, Y., Luo, T., Zhu, Z., Shan, Y., Matoba, N., et al. (2022). Common variants contribute to intrinsic human brain functional networks. Nature Genetics, 54(4), 508–517.

Zhao, B., Li, T., Yang, Y., Wang, X., Luo, T., Shan, Y., Zhu, Z., Xiong, D., Hauberg, M. E., Bendl, J., et al. (2021). Common genetic variation influencing human white matter microstructure. Science, 372(6548), eabf3736.

Zhao, Y., Chen, T., Cai, J., Lichenstein, S., Potenza, M. N., & Yip, S. W. (2022). Bayesian network mediation analysis with application to the brain functional connectome. Statistics in Medicine, 41(20), 3991– 4005.

Zhou, X., Tang, W., Greenwood, T. A., Guo, S., He, L., Geyer, M. A., & Kelsoe, J. R. (2009). Transcription factor sp4 is a susceptibility gene for bipolar disorder. PloS One, 4(4), e5196.

