## Supplemental Materials for "The Mediating Role of Structural Connectivity in Genetic Effects on Functional Brain Networks"

### Supplementary Material

The supplementary materials contain:

1. Figures S1 and S2, which complement Section 3.1.1.

2. Tables S1 and S2, corresponding to Section 2.2 and Section 2.3.

3. Tables S3–S6, presenting results related to Section 3.3.2.

4. A table of significant indirect effect pathways, included with the initial submission.

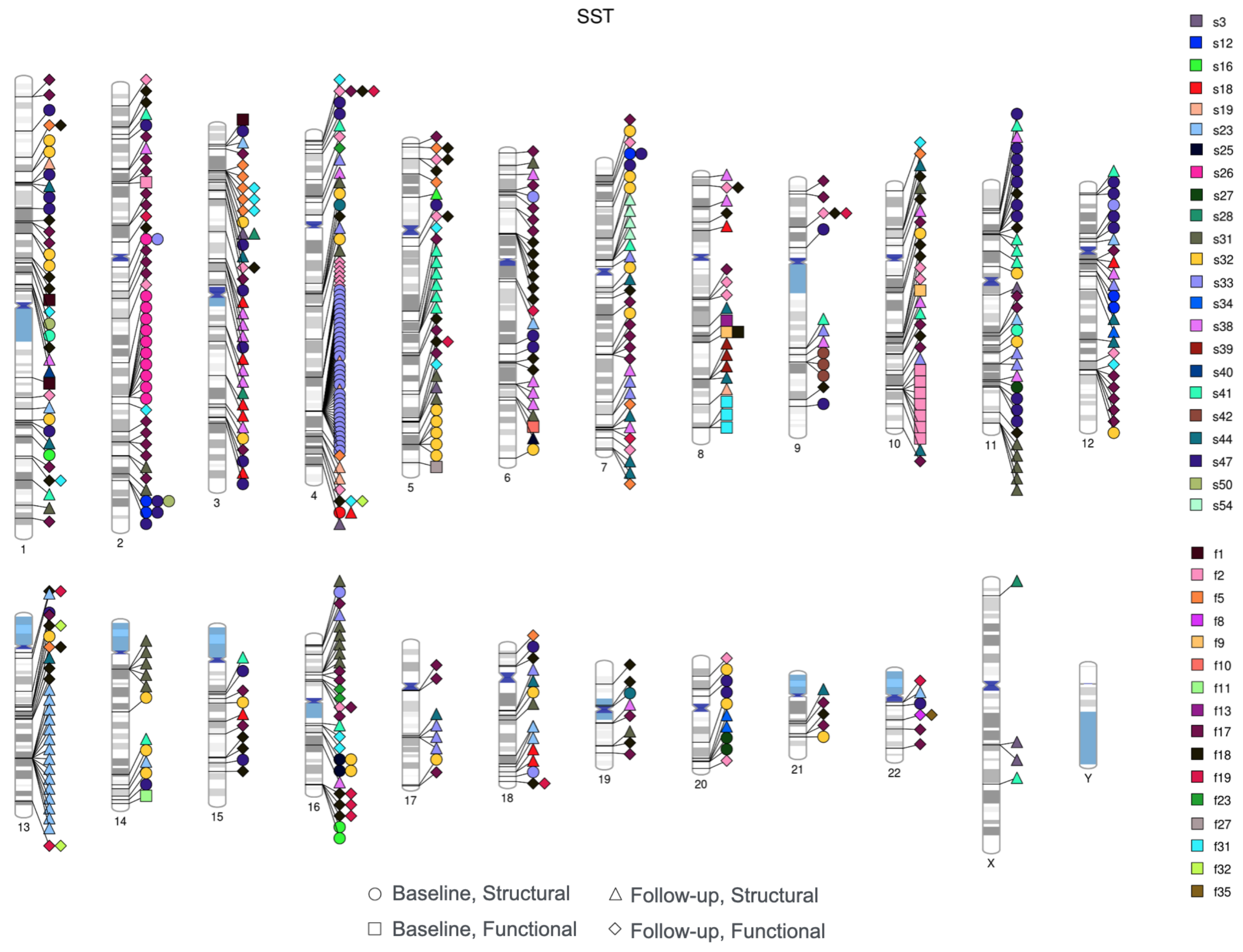


Figure S1: Ideogram of genomic Network Traitss for SST.

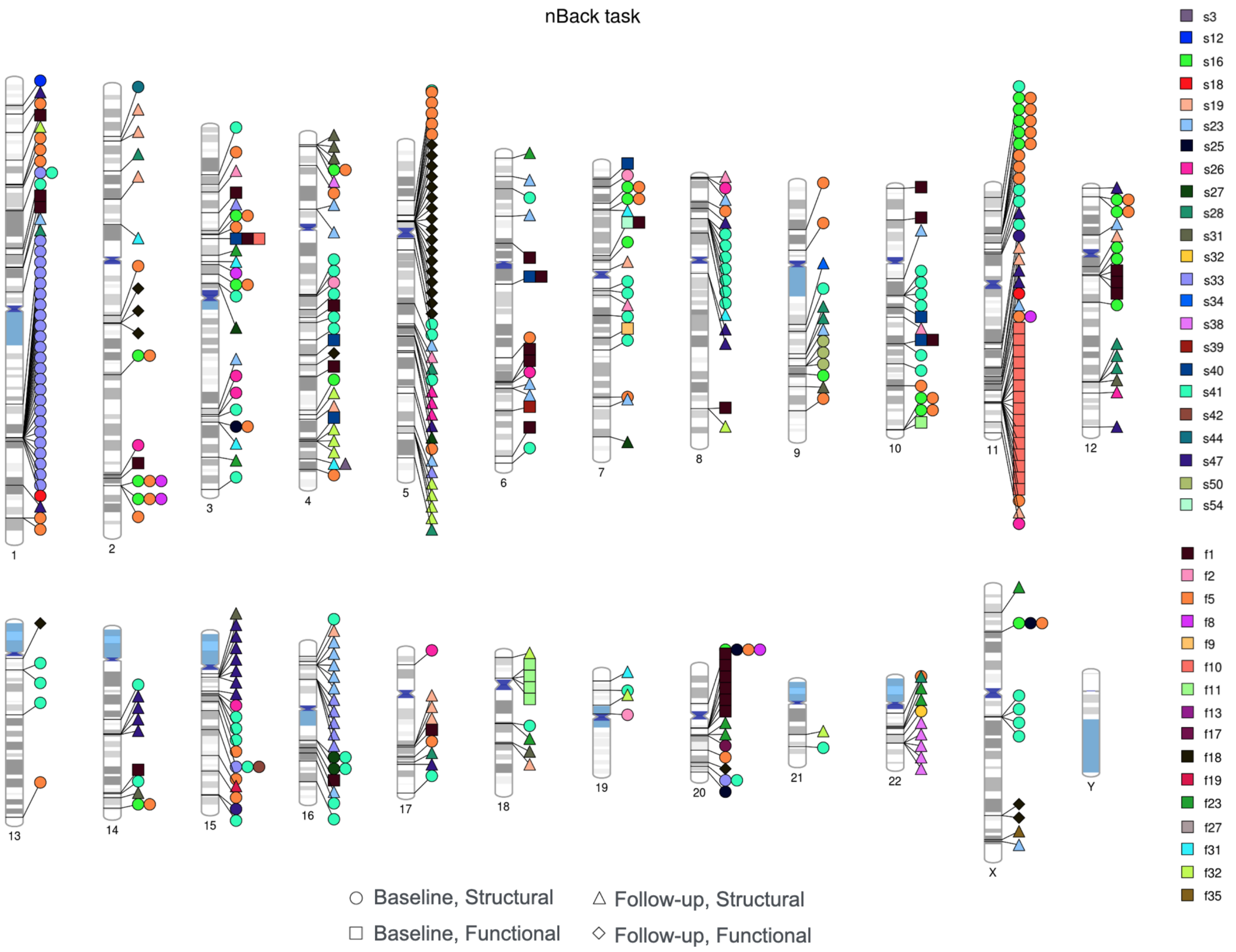


Figure S2: Ideogram of genomic Network Traitss for nBack task.

Table S1: Symbols corresponding to functional network traits.

| Symbol | Network Traits | Symbol | Network Traits |
|---|---|---|---|
| f1 | (Medial frontal,Medial frontal) | f19 | (Frontoparietal,Visual I) |
| f2 | (Frontoparietal,Frontoparietal) | f20 | (Frontoparietal,Visual II) |
| f3 | (DMN,DMN) | f21 | (Frontoparietal,Visual association) |
| f4 | (SC-Cer,SC-Cer) | f22 | (DMN,SC-Cer) |
| f5 | (Motor,Motor) | f23 | (DMN,Motor) |
| f6 | (Visual I,Visual I) | f24 | (DMN,Visual I) |
| f7 | (Visual II,Visual II) | f25 | (DMN,Visual II) |
| f8 | (Visual association,Visual association) | f26 | (DMN,Visual association) |
| f9 | (Medial frontal,Frontoparietal) | f27 | (SC-Cer,Motor) |
| f10 | (Medial frontal,DMN) | f28 | (SC-Cer,Visual I) |
| f11 | (Medial frontal,SC-Cer) | f29 | (SC-Cer,Visual II) |
| f12 | (Medial frontal,Motor) | f30 | (SC-Cer,Visual association) |
| f13 | (Medial frontal,Visual I) | f31 | (Motor,Visual I) |
| f14 | (Medial frontal,Visual II) | f32 | (Motor,Visual II) |
| f15 | (Medial frontal,Visual association) | f33 | (Motor,Visual association) |
| f16 | (Frontoparietal,DMN) | f34 | (Visual I,Visual II) |
| f17 | (Frontoparietal,SC-Cer) | f35 | (Visual I,Visual association) |
| f18 | (Frontoparietal,Motor) | f36 | (Visual II,Visual association) |

Table S2: Symbols corresponding to structural network traits.

| Symbol | Network Traits | Symbol | Network Traits |
|---|---|---|---|
| s1 | (lh.frontal,lh.frontal) | s29 | (lh.limbic,lh.occipital) |
| s2 | (lh.parietal,lh.parietal) | s30 | (lh.limbic,rh.frontal) |
| s3 | (lh.limbic,lh.limbic) | s31 | (lh.limbic,rh.parietal) |
| s4 | (lh.temporal,lh.temporal) | s32 | (lh.limbic,rh.limbic) |
| s5 | (lh.occipital,lh.occipital) | s33 | (lh.limbic,rh.temporal) |
| s6 | (rh.frontal,rh.frontal) | s34 | (lh.limbic,rh.occipital) |
| s7 | (rh.parietal,rh.parietal) | s35 | (lh.temporal,lh.occipital) |
| s8 | (rh.limbic,rh.limbic) | s36 | (lh.temporal,rh.frontal) |
| s9 | (rh.temporal,rh.temporal) | s37 | (lh.temporal,rh.parietal) |
| s10 | (rh.occipital,rh.occipital) | s38 | (lh.temporal,rh.limbic) |
| s11 | (lh.frontal,lh.parietal) | s39 | (lh.temporal,rh.temporal) |
| s12 | (lh.frontal,lh.limbic) | s40 | (lh.temporal,rh.occipital) |
| s13 | (lh.frontal,lh.temporal) | s41 | (lh.occipital,rh.frontal) |
| s14 | (lh.frontal,lh.occipital) | s42 | (lh.occipital,rh.parietal) |
| s15 | (lh.frontal,rh.frontal) | s43 | (lh.occipital,rh.limbic) |
| s16 | (lh.frontal,rh.parietal) | s44 | (lh.occipital,rh.temporal) |
| s17 | (lh.frontal,rh.limbic) | s45 | (lh.occipital,rh.occipital) |
| s18 | (lh.frontal,rh.temporal) | s46 | (rh.frontal,rh.parietal) |
| s19 | (lh.frontal,rh.occipital) | s47 | (rh.frontal,rh.limbic) |
| s20 | (lh.parietal,lh.limbic) | s48 | (rh.frontal,rh.temporal) |
| s21 | (lh.parietal,lh.temporal) | s49 | (rh.frontal,rh.occipital) |
| s22 | (lh.parietal,lh.occipital) | s50 | (rh.parietal,rh.limbic) |
| s23 | (lh.parietal,rh.frontal) | s51 | (rh.parietal,rh.temporal) |
| s24 | (lh.parietal,rh.parietal) | s52 | (rh.parietal,rh.occipital) |
| s25 | (lh.parietal,rh.limbic) | s53 | (rh.limbic,rh.temporal) |
| s26 | (lh.parietal,rh.temporal) | s54 | (rh.limbic,rh.occipital) |
| s27 | (lh.parietal,rh.occipital) | s55 | (rh.temporal,rh.occipital) |
| s28 | (lh.limbic,lh.temporal) | | |

Table S3: A full list of mediation pathways for eQTLs identified for the resting-state condition.

| SNP | Cytoband | p-value | Regulated genes | Functional network | Structural network |
|---|---|---|---|---|---|
| rs144067396 | p11.32 | 2.71E-05 | SMCHD1 | (SC-Cer,SC-Cer) | (lh.frontal,lh.parietal) |
| rs74101672 | q31.1 | 2.57E-05 | SLITRK6 | (Visual association, Visual association) | (lh.occipital,rh.occipital) |
| rs143925313 | q13.13 | 2.08E-05 | ADNP | (Frontoparietal,Visual I) | (lh.frontal,lh.frontal) |
| rs143925313 | q13.13 | 1.27E-06 | DPM1 | (Frontoparietal,Visual I) | (lh.frontal,lh.frontal) |

Table S4: A full list of mediation pathways for eQTLs identified for the MID task.

| SNP | Cytoband | p-value | Regulated genes | Functional network | Structural network |
|---|---|---|---|---|---|
| rs1407504 | q22.33 | 1.33E-05 | COL15A1 | (Frontoparietal,Frontoparietal) | (lh.frontal,rh.frontal) |
| rs1407504 | q22.33 | 1.33E-05 | COL15A1 | (DMN,DMN) | (lh.frontal,rh.frontal) |
| rs1407504 | q22.33 | 1.33E-05 | COL15A1 | (SC-Cer,SC-Cer) | (lh.frontal,rh.frontal) |
| rs1407504 | q22.33 | 1.33E-05 | COL15A1 | (Visual I,Visual I) | (lh.frontal,rh.frontal) |
| rs1407504 | q22.33 | 1.33E-05 | COL15A1 | (Visual II,Visual II) | (lh.frontal,rh.frontal) |
| rs1407504 | q22.33 | 1.33E-05 | COL15A1 | (Visual association, Visual association) | (lh.frontal,rh.frontal) |
| rs1407504 | q22.33 | 1.33E-05 | COL15A1 | (Visual association, Visual association) | (lh.occipital,rh.occipital) |
| rs1407504 | q22.33 | 1.33E-05 | COL15A1 | (Frontoparietal,SC-Cer) | (lh.frontal,rh.frontal) |
| rs1407504 | q22.33 | 1.33E-05 | COL15A1 | (Frontoparietal,Visual I) | (lh.frontal,rh.frontal) |
| rs1407504 | q22.33 | 1.33E-05 | COL15A1 | (Frontoparietal, Visual association) | (lh.occipital,rh.occipital) |
| rs1407504 | q22.33 | 1.33E-05 | COL15A1 | (DMN,Visual I) | (lh.frontal,rh.frontal) |
| rs1407504 | q22.33 | 1.33E-05 | COL15A1 | (Motor,Visual I) | (lh.frontal,lh.frontal) |
| rs1407504 | q22.33 | 1.33E-05 | COL15A1 | (Motor,Visual I) | (lh.frontal,rh.frontal) |
| rs1407504 | q22.33 | 1.33E-05 | COL15A1 | (Visual I,Visual II) | (lh.frontal,rh.frontal) |
| rs1407504 | q22.33 | 1.33E-05 | COL15A1 | (Visual II, Visual association) | (lh.frontal,rh.frontal) |

Table S5: A full list of mediation pathways for eQTLs identified for the SST.

| SNP | Cytoband | p-value | Regulatedgenes | Functional Subnetworks | Structural Subnetworks |
|---|---|---|---|---|---|
| rs58977077 | q31.3 | 1.31E-15 | SH3D19 | (SC-Cer,SC-Cer) | (lh.parietal,rh.parietal) |
| rs58977077 | q31.3 | 4.24E-23 | FAM160A1 | (SC-Cer,SC-Cer) | (lh.parietal,rh.parietal) |
| rs58977077 | q31.3 | 5.34E-22 | PET112 | (SC-Cer,SC-Cer) | (lh.parietal,rh.parietal) |
| rs58977077 | q31.3 | 1.31E-15 | SH3D19 | (Motor,Motor) | (lh.parietal,rh.parietal) |
| rs58977077 | q31.3 | 4.24E-23 | FAM160A1 | (Motor,Motor) | (lh.parietal,rh.parietal) |
| rs58977077 | q31.3 | 5.34E-22 | PET112 | (Motor,Motor) | (lh.parietal,rh.parietal) |
| rs58977077 | q31.3 | 1.31E-15 | SH3D19 | (Motor,Motor) | (rh.frontal,rh.limbic) |
| rs58977077 | q31.3 | 4.24E-23 | FAM160A1 | (Motor,Motor) | (rh.frontal,rh.limbic) |
| rs58977077 | q31.3 | 5.34E-22 | PET112 | (Motor,Motor) | (rh.frontal,rh.limbic) |
| rs58977077 | q31.3 | 1.31E-15 | SH3D19 | (Visual I,Visual I) | (lh.parietal,rh.parietal) |
| rs58977077 | q31.3 | 4.24E-23 | FAM160A1 | (Visual I,Visual I) | (lh.parietal,rh.parietal) |
| rs58977077 | q31.3 | 5.34E-22 | PET112 | (Visual I,Visual I) | (lh.parietal,rh.parietal) |
| rs58977077 | q31.3 | 1.31E-15 | SH3D19 | (Visual association, Visual association) | (lh.parietal,rh.parietal) |
| rs58977077 | q31.3 | 4.24E-23 | FAM160A1 | (Visual association, Visual association) | (lh.parietal,rh.parietal) |
| rs58977077 | q31.3 | 5.34E-22 | PET112 | (Visual association, Visual association) | (lh.parietal,rh.parietal) |
| rs58977077 | q31.3 | 1.31E-15 | SH3D19 | (Medial frontal,Visual I) | (lh.parietal,rh.parietal) |
| rs58977077 | q31.3 | 4.24E-23 | FAM160A1 | (Medial frontal,Visual I) | (lh.parietal,rh.parietal) |
| rs58977077 | q31.3 | 5.34E-22 | PET112 | (Medial frontal,Visual I) | (lh.parietal,rh.parietal) |
| rs58977077 | q31.3 | 1.31E-15 | SH3D19 | (DMN,SC-Cer) | (lh.parietal,rh.parietal) |
| rs58977077 | q31.3 | 4.24E-23 | FAM160A1 | (DMN,SC-Cer) | (lh.parietal,rh.parietal) |
| rs58977077 | q31.3 | 5.34E-22 | PET112 | (DMN,SC-Cer) | (lh.parietal,rh.parietal) |
| rs58977077 | q31.3 | 1.31E-15 | SH3D19 | (DMN,Visual I) | (lh.parietal,rh.parietal) |
| rs58977077 | q31.3 | 4.24E-23 | FAM160A1 | (DMN,Visual I) | (lh.parietal,rh.parietal) |
| rs58977077 | q31.3 | 5.34E-22 | PET112 | (DMN,Visual I) | (lh.parietal,rh.parietal) |
| rs58977077 | q31.3 | 1.31E-15 | SH3D19 | (DMN,Visual II) | (lh.parietal,rh.parietal) |
| rs58977077 | q31.3 | 4.24E-23 | FAM160A1 | (DMN,Visual II) | (lh.parietal,rh.parietal) |
| rs58977077 | q31.3 | 5.34E-22 | PET112 | (DMN,Visual II) | (lh.parietal,rh.parietal) |
| rs58977077 | q31.3 | 1.31E-15 | SH3D19 | (SC-Cer,Visual I) | (lh.parietal,rh.parietal) |
| rs58977077 | q31.3 | 4.24E-23 | FAM160A1 | (SC-Cer,Visual I) | (lh.parietal,rh.parietal) |
| rs58977077 | q31.3 | 5.34E-22 | PET112 | (SC-Cer,Visual I) | (lh.parietal,rh.parietal) |
| rs58977077 | q31.3 | 1.31E-15 | SH3D19 | (SC-Cer, Visual association) | (lh.parietal,rh.parietal) |
| rs58977077 | q31.3 | 4.24E-23 | FAM160A1 | (SC-Cer, Visual association) | (lh.parietal,rh.parietal) |
| rs58977077 | q31.3 | 5.34E-22 | PET112 | (SC-Cer, Visual association) | (lh.parietal,rh.parietal) |
| rs58977077 | q31.3 | 1.31E-15 | SH3D19 | (Motor,Visual I) | (rh.frontal,rh.limbic) |
| rs58977077 | q31.3 | 4.24E-23 | FAM160A1 | (Motor,Visual I) | (rh.frontal,rh.limbic) |
| rs58977077 | q31.3 | 5.34E-22 | PET112 | (Motor,Visual I) | (rh.frontal,rh.limbic) |
| rs58977077 | q31.3 | 1.31E-15 | SH3D19 | (Motor, Visual association) | (lh.parietal,rh.parietal) |
| rs58977077 | q31.3 | 4.24E-23 | FAM160A1 | (Motor, Visual association) | (lh.parietal,rh.parietal) |
| rs58977077 | q31.3 | 5.34E-22 | PET112 | (Motor, Visual association) | (lh.parietal,rh.parietal) |
| rs58977077 | q31.3 | 1.31E-15 | SH3D19 | (Visual I,Visual II) | (lh.parietal,rh.parietal) |
| rs58977077 | q31.3 | 4.24E-23 | FAM160A1 | (Visual I,Visual II) | (lh.parietal,rh.parietal) |
| rs58977077 | q31.3 | 5.34E-22 | PET112 | (Visual I,Visual II) | (lh.parietal,rh.parietal) |
| rs58977077 | q31.3 | 1.31E-15 | SH3D19 | (Visual I, Visual association) | (lh.parietal,rh.parietal) |
| rs58977077 | q31.3 | 4.24E-23 | FAM160A1 | (Visual I, Visual association) | (lh.parietal,rh.parietal) |
| rs58977077 | q31.3 | 5.34E-22 | PET112 | (Visual I, Visual association) | (lh.parietal,rh.parietal) |

Table S6: A full list of mediation pathways for eQTLs identified for the nBack task.

| SNP | Cytoband | p-value | Regulated genes | Functional networks | Structural networks |
|---|---|---|---|---|---|
| rs6060311 | q11.22 | 3.19E-05 | NCOA6 | (SC-Cer,SC-Cer) | (lh.parietal,rh.temporal) |
| rs6060311 | q11.22 | 1.18E-04 | CEP250 | (SC-Cer,SC-Cer) | (lh.parietal,rh.temporal) |
| rs6060311 | q11.22 | 3.19E-05 | NCOA6 | (SC-Cer,SC-Cer) | (lh.temporal,rh.parietal) |
| rs6060311 | q11.22 | 1.18E-04 | CEP250 | (SC-Cer,SC-Cer) | (lh.temporal,rh.parietal) |
| rs6060311 | q11.22 | 3.19E-05 | NCOA6 | (Visual I,Visual I) | (lh.parietal,rh.temporal) |
| rs6060311 | q11.22 | 1.18E-04 | CEP250 | (Visual I,Visual I) | (lh.parietal,rh.temporal) |
| rs141217457 | q15.3 | 1.68E-05 | CATSPER2 | (Visual association, Visual association) | (rh.frontal,rh.occipital) |
| rs141217457 | q15.3 | 1.68E-05 | CATSPER2 | (Frontoparietal,SC-Cer) | (rh.frontal,rh.occipital) |
| rs141217457 | q15.3 | 1.68E-05 | CATSPER2 | (Frontoparietal,Visual I) | (rh.frontal,rh.occipital) |
| rs6060311 | q11.22 | 3.19E-05 | NCOA6 | (DMN,Motor) | (lh.parietal,rh.temporal) |
| rs6060311 | q11.22 | 1.18E-04 | CEP250 | (DMN,Motor) | (lh.parietal,rh.temporal) |
| rs141217457 | q15.3 | 1.68E-05 | CATSPER2 | (SC-Cer,Motor) | (rh.frontal,rh.occipital) |
| rs6060311 | q11.22 | 3.19E-05 | NCOA6 | (SC-Cer,Visual I) | (lh.parietal,rh.temporal) |
| rs6060311 | q11.22 | 1.18E-04 | CEP250 | (SC-Cer,Visual I) | (lh.parietal,rh.temporal) |
| rs6060311 | q11.22 | 3.19E-05 | NCOA6 | (SC-Cer,Visual I) | (lh.temporal,rh.temporal) |
| rs6060311 | q11.22 | 1.18E-04 | CEP250 | (SC-Cer,Visual I) | (lh.temporal,rh.temporal) |
| rs141217457 | q15.3 | 1.68E-05 | CATSPER2 | (SC-Cer,Visual I) | (rh.frontal,rh.occipital) |
| rs6060311 | q11.22 | 3.19E-05 | NCOA6 | (SC-Cer, Visual association) | (lh.parietal,rh.temporal) |
| rs6060311 | q11.22 | 1.18E-04 | CEP250 | (SC-Cer, Visual association) | (lh.parietal,rh.temporal) |
| rs6060311 | q11.22 | 3.19E-05 | NCOA6 | (SC-Cer, Visual association) | (lh.temporal,rh.parietal) |
| rs6060311 | q11.22 | 1.18E-04 | CEP250 | (SC-Cer, Visual association) | (lh.temporal,rh.parietal) |
| rs141217457 | q15.3 | 1.68E-05 | CATSPER2 | (Motor,Visual I) | (rh.frontal,rh.occipital) |
| rs6060311 | q11.22 | 3.19E-05 | NCOA6 | (Visual I, Visual association) | (lh.parietal,rh.temporal) |
| rs6060311 | q11.22 | 1.18E-04 | CEP250 | (Visual I, Visual association) | (lh.parietal,rh.temporal) |
| rs6060311 | q11.22 | 3.19E-05 | NCOA6 | (Visual I, Visual association) | (lh.temporal,rh.temporal) |
| rs6060311 | q11.22 | 1.18E-04 | CEP250 | (Visual I, Visual association) | (lh.temporal,rh.temporal) |
| rs141217457 | q15.3 | 1.68E-05 | CATSPER2 | (Visual I, Visual association) | (rh.frontal,rh.occipital) |
